# Comprehensive Measurement of Inter-Individual Variation in DNA Repair Capacity in Healthy Individuals

**DOI:** 10.1101/2025.06.13.25329369

**Authors:** Ting Zhai, Patrizia Mazzucato, Catherine Ricciardi, David C. Christiani, Liming Liang, Leona D. Samson, Isaac A. Chaim, Zachary D. Nagel

## Abstract

Rare genetic DNA repair deficiency syndromes can lead to immunodeficiency, neurological disorders, and cancer. In the general population, inter-individual variation in DNA repair capacity (DRC) influences susceptibility to cancer and several age-related diseases. Genome wide association studies and functional analyses show that defects in multiple DNA repair pathways jointly increase disease risk, but previous technologies did not permit comprehensive analyses of DNA repair in populations. To overcome these limitations, we used fluorescence multiplex host cell reactivation (FM-HCR) assays that directly quantify DRC across six major DNA repair pathways. We assessed DRC in phytohemagglutinin-stimulated primary lymphocytes from 56 healthy individuals and validated assay reproducibility in 10 individuals with up to five independent blood draws. We furthermore developed generalized analytical pipelines for systematically adjusting for batch effects and both experimental and biological confounders. Our results reveal significant inter-individual variation in DRC for each of 10 reporter assays that measure the efficiency of distinct repair processes. Our data also demonstrate that correlations between the activities of different DNA repair pathways are relatively weak. This finding suggests that each pathway may independently influence susceptibility to the health effects of DNA damage. We furthermore developed a pipeline for analyzing comet repair kinetics and related our new functional data to previously reported comet assay data for the same individuals.

Our pioneering analysis underscores the sensitivity of FM-HCR assays for detecting subtle biological differences between individuals and establishes standardized methodologies for population studies. Our findings and open source analytical tools advance precision medicine by enabling comprehensive exploration of genetic, demographic, clinical, and lifestyle factors and supporting targeted interventions to enhance DNA repair and maintain genomic integrity, thereby promoting personalized healthcare and disease prevention.

## Introduction

A longstanding goal of precision medicine is to identify individuals at high risk of disease, enabling early detection and personalized prevention ^1^. DNA repair deficiency can lead to cancer, immune disorders, neurological disease, and premature aging ^2–4^. Assessing individuals’ DNA repair capacity (DRC) against damage in human populations, therefore, holds great potential for facilitating risk stratification and targeted interventions for many diseases ^5^; accumulating evidence supports this paradigm, but progress has been hampered by limitations of available technologies.

Extensive population studies, including genome-wide association studies (GWAS), have demonstrated that germline variation in genes from multiple DNA repair pathways contribute to disease susceptibility ^6,7^. However, GWAS rely on hereditary information at the population level, often requiring large sample sizes and the combined effects of numerous variants to achieve statistical significance in predictive models. This approach may not provide actionable insights at the individual level. Additionally, environmental factors such as exposure to DNA-damaging agents and aging influence an individual’s DRC ^8^. DRC can therefore be dynamic over the lifespan and lead to environmentally determined inter-individual variability that is not captured by genetic data alone ^9^.

Somatic mutation signatures, which integrate the biological effects of both exposure to DNA damaging agents and defects in DNA repair have also emerged as a strategy for assessing genome maintenance ^10,11^. However somatic mutation signatures that are characteristic of specific types of DNA damage or DNA repair deficiencies ^10^ reflect the cumulative consequences of genome instability and thus may not reflect the current status of DRC ^11^. Furthermore, mutation signatures and other biomarkers derived from large-scale multi-omics sequencing data in tumors ^12^ may not be applicable to non-cancerous cells that have more homogeneous omics profiles and in populations with less frequent exposures. They are also subject to sequencing noise and may be less sensitive due to computational challenges in accurately detecting and interpreting rare variants or low-abundance signals ^13^. Finally, biomarkers established from tumor samples may not translate well to the general population due to the unique genetics and microenvironment of tumors.

Functional assays can provide accurate, individualized, real-time measures of DRC that are more powerful predictors of cancer risk than genetic scores, but population studies have almost exclusively included only one DNA repair pathway at a time ^14–17^. Host-cell reactivation (HCR) assays functionally assess DRC by measuring the repair of plasmids modified with DNA damage that affects the expression of a reporter gene. Originally applied in population studies by Grossman and Wei ^18,19^ and later expanded by Spitz ^20^, David ^21^, and others ^22–28^, HCR assays integrate complex layers of cellular regulation, including genetic variants, epigenetics, transcription, and translation, into a single readout. Traditional HCR assays, however, have not been extensively applied population-level studies due to limitations in throughput and concerns about assay variability. They target individual repair pathways and consequently require a large amount of biological material for scaling assays across multiple pathways ^9^. They have also historically been limited to DNA lesions that either blocked transcription or led to a change in the sequence of the repaired DNA to produce a signal. In addition to these assay limitations, a systematic strategy for addressing missing values, technical variability, and batch effects that are critical in population studies has been lacking.

To address the assay-dependent limitations, we previously developed fluorescence multiplex host cell reactivation (FM-HCR) assays, which enable simultaneous measurements of repair capacity in all major DNA repair pathways ^29–31^. The use of transient transfection sets the FM-HCR approach apart from assays that require stable integration of reporter systems into the genomic DNA and are thus incompatible with analyses in primary cells, populations, and most types of DNA damage. Importantly, measurements in the context of live cells ensure that the repair machinery is assessed within its physiological environment, making FM-HCR assays a powerful tool for evaluating DRC at the individual level. The multiplex design reduces the required sample input, facilitating large-scale population studies by allowing multiple pathways to be assessed concurrently using highly sensitive flow cytometric quantitation of reporter protein expression. While FM-HCR assays have been used in PBMCs, the assay’s accuracy and reproducibility at the population level has not been rigorously tested.

In this study, we evaluate the applicability of the FM-HCR assay for population-level research by comparing the activity of six major DNA repair pathways in primary human lymphocytes from a cohort of 56 healthy individuals, including repeated measurements in a subset of donors. We furthermore introduce an analytical pipeline that systematically controls for technical variability and corrects for batch effects inherent to population-based data. This dataset adds to previous population-scale characterizations of nucleotide excision repair (NER), base excision repair (BER), non-homologous end joining (NHEJ) and direct reversal by the MGMT protein. The data also provide insights into how mismatch repair (MMR), homologous recombination (HR), and MUTYH-dependent initiation of A:8oxoG repair, vary among individuals and within-person over time. We also correlate our FM-HCR measurements between DNA repair pathways with repair kinetics for oxidative DNA damage measured and reported previously using the CometChip assay in the same cohort ^32^. Our findings demonstrate the FM-HCR assay’s potential to provide accurate, individualized assessments of DRC, thereby enhancing risk stratification and informing personalized preventative and therapeutic strategies in precision medicine.

## Results

Among 56 enrolled participants, 10 provided repeated measurements with 4 or 5 blood draws over 4-6 weeks (**Fig. 1**). Participants ages spanned from 20-66 years, with a median of 28 years and 55.4% female representation (**Table 1**). The majority of participants were white (73.2%) and never smokers (82.1%), and the median BMI was 22.76 kg/m^2^. FM-HCR assays were conducted in triplicates per individual per blood draw, contributing to a total of 285 measurements x 10 assays that were measured across 37 batches. The range of repair capacity (represented by reporter expression %) varied from approximately 1.5-fold (NHEJ, 8oxoG:C, long-patch [LP]-BER) to over 10-fold (A:8oxoG, MGMT) across the 56 individuals at their initial visits (**Fig. 2**).

**Fig. 1.**
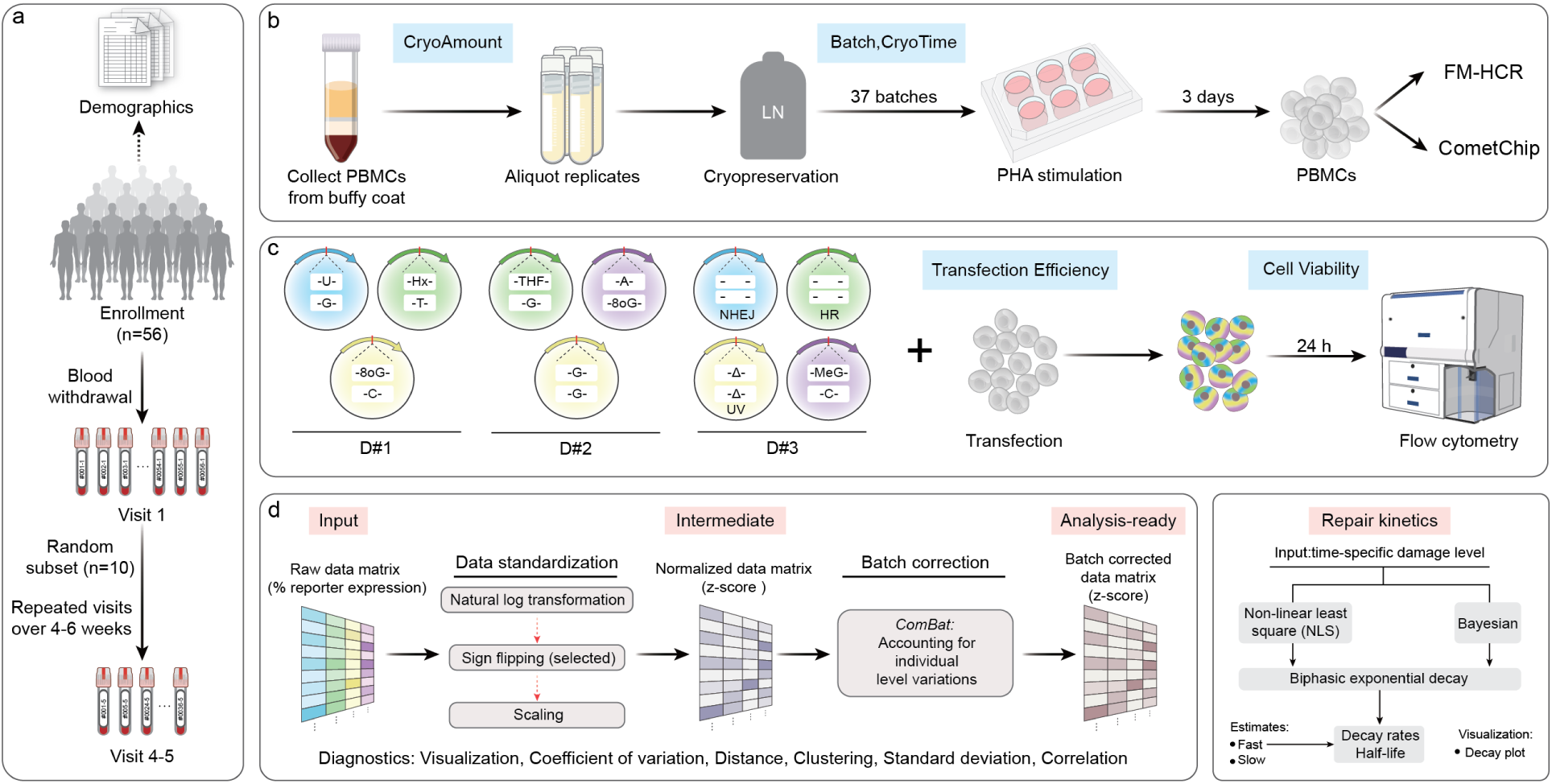
Study design and workflow. **(a)** Study design: Demographic data were collected at baseline using a structured questionnaire. Blood samples were collected from 56 apparently healthy individuals during their first visit. A subset of 10 participants returned for 4-5 additional blood draws over a period of 4-6 weeks. **(b)** Blood sample processing: Peripheral blood mononuclear cells (PBMCs) were isolated from whole blood using Ficoll density gradient centrifugation, cryopreserved in liquid nitrogen, and subsequently thawed and stimulated with phytohemagglutinin-L (PHA-L) for three days. Stimulated PBMCs were aliquoted for downstream assays including DNA repair capacity measurement using fluorescence multiplex host cell reactivation (FM-HCR) assays, and DNA repair kinetics assessment by the CometChip assay following hydrogen peroxide (H2O2) treatment. Technical variables potentially impacting genome integrity assay measurements were also recorded. **(c)** FM-HCR workflow: DNA damage containing reporter plasmids were transiently transfected into stimulated lymphocytes with by electroporating 3 separate cocktails of 10 fluorescent reporter plasmids (D#1, D#2, D#3), each containing a specific type of DNA damage, or cocktails containing damage free plasmids (UD#1, UD#2, not shown). Repair efficiency was determined based on the expression levels of fluorescent reporter genes, quantified via flow cytometry. **(d)** Data processing pipeline: We developed a custom R package and web interface to streamline the processing and analysis of DNA repair data at the population level. For FM-HCR, the pipeline standardizes raw data, performs batch correction, and outputs analysis-ready data, with diagnostics to evaluate processing efficiency. For CometChip, the pipeline analyzes repair kinetics using non-linear least squares or Bayesian approaches to fitting a biphasic exponential decay model, and provides estimates of overall, fast-and slow-phase half-lives. It also automatically generates sample-specific decay plots for more detailed visualization.

**Fig. 2.**
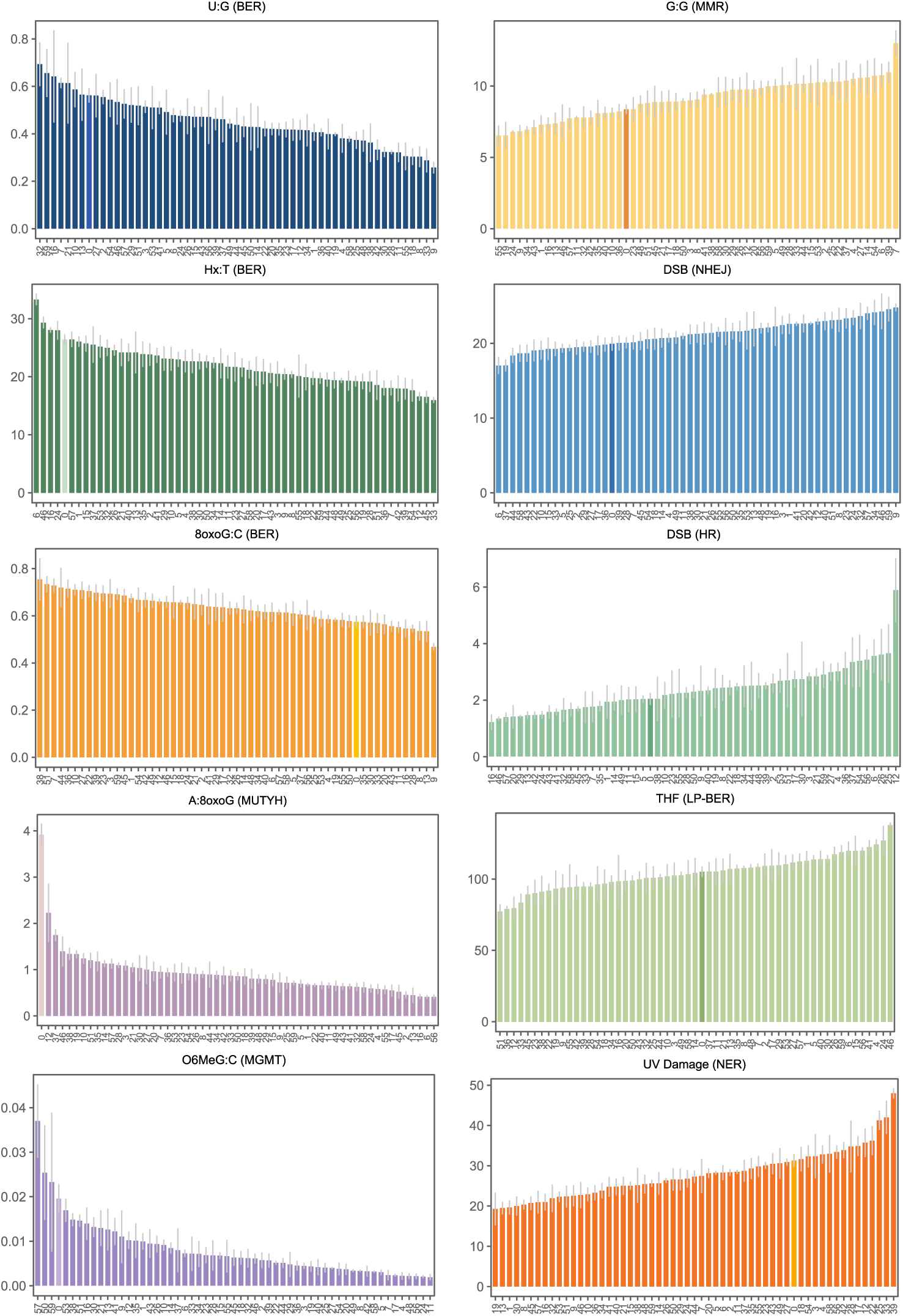
Inter-individual variation in reporter expression across 10 substrates. Each bar represents the mean of three technical replicates for a single individual (labeled with a participant number between 1 and 59), with error bars indicating the standard error. A control PBMC sample that was included in every batch is highlighted in a distinct color and assigned a participant number “0”. Individuals are ordered from lowest to highest repair capacity within each pathway. Data were batch-corrected and reverted to the raw reporter expression scale. Data for the control PBMC sample were not corrected, since they were included in each batch and therefore the mean of all 37 control replicates already reflect the overall batch average.

**Table 1.** Demographics of study subjects.

| Variable | Summary |
| --- | --- |
| Age (years, median [IQR]) | 28 (25-34) |
| Sex (female %) | 31 (55.4%) |
| Race |  |
| White | 41 (73.2%) |
| Asian | 12 (21.4%) |
| Black/African American | 3 (0.05%) |
| Weight (lb, median [IQR]) | 149 (130-173) |
| Height (cm, median [IQR]) | 172 (168-178) |
| BMI (kg/m<sup>2</sup>, median [IQR]) | 22.8 (20.9-25.5) |
| Smoking status |  |
| Never | 46 (82.1%) |
| Former | 8 (14.3%) |
| Current | 2 (0.04%) |
| Place of birth (US %) | 36 (64.3%) |
| Family history of cancer (yes %) | 33 (58.9%) |
| Alcohol consumption (ever %) | 50 (89.3%) |
| CryoAmount (millions per tube) | 13.7 (11.8-17.4) |
| CryoTime (days) | 88 (75-87) |
| Cell viability | 29.6 (21.4-35.2) |
| Transfection efficiency | 15.8 (12.1-19.7) |

IQR: inter-quantile range; BMI: body mass index; CryoAmount: cell amount available for cryopreservation after isolation; CryoTime: duration of cryopreservation.

### Controlling technical variability improves FM-HCR reproducibility

We first explored the influences of several key technical and biological factors arising from sample processing and experimental conditions, based on our previous applications of FM-HCR. These included batch, cryopreservation time (CryoTime), cell amount from blood isolation (CryoAmount), cell viability, and transfection efficiency (TE), as detailed in **Supplementary methods**. The coefficient of variation (CV) was calculated to quantify variability, which better accounted for sample size differences and outliers than fold-change. Batch effects emerged as the primary source of variation in the raw FM-HCR readouts, with across-batch CVs ranging from 12.4% to 45.7% and within-individual CVs ranging from 15.9% to 46.5% (**Supplementary** Fig. 1-2), highlighting the need for adjustment before true inter-individual variability can be reliably distinguished.

Using our tailored analytical strategy (**Supplementary methods**), we substantially reduced batch-related variability, with the across-batch CV reduced by an average of 74.0% and the within-individual CV by 31.5% (**Supplementary Table 1**).

Moreover, repeated measures from the same individual became more clustered, with the average CV decreasing from 20.1% to 16.7%, suggesting improved reproducibility (**Supplementary Table 2**). Mixed technical-biological variables (CryoAmount, cell viability, and TE) retained modest, pathway-specific associations with DRC after adjustment (**Supplementary methods; Supplementary** Fig. 3-4), further explaining the remaining within-individual variation (**Supplementary discussion**). These findings underscore the importance of consistent sample handling and inclusion of sample-specific technical and biological parameters in statistical models.

### Inter-individual variation in DNA repair capacity

Significant inter-individual variation was detected across the 10 reporter assays (**Table 2**). A landscape of relative DRC is presented in **Fig. 3a**, along with demographics. Probing the factors contributing to the observed inter-individual variation, we discovered that demographic factors including age, race, and smoking status all had independent roles in influencing repair capacity in one or more pathways (**Fig. 4**). We observed that A:8oxoG repair capacity decreased with age (est. decrease of 0.02 standard deviations per year, p = 0.004 [false discovery rate, FDR = 0.037]). Race was associated with differences in DRC in several pathways including Hx:T, A:8oxoG, MMR, NHEJ, MGMT, and NER. After multiple-testing correction, the associations remained statistically significant for Hx:T, A:8oxoG, MMR, and NER. And smoking was associated with observed decreased repair in LP-BER (current vs never smoker: est.= −0.83 standard deviations, p = 0.026 [FDR = 0.258]), though the significance diminished after multiple-testing adjustment. To further ensure the robustness of our findings, we conducted a sensitivity analysis using complete case data. The results were consistent with those obtained through multiple imputation, indicating that the observed associations were not substantially affected by the imputation of missing values (**Supplementary Table 3**).

**Fig. 3.**
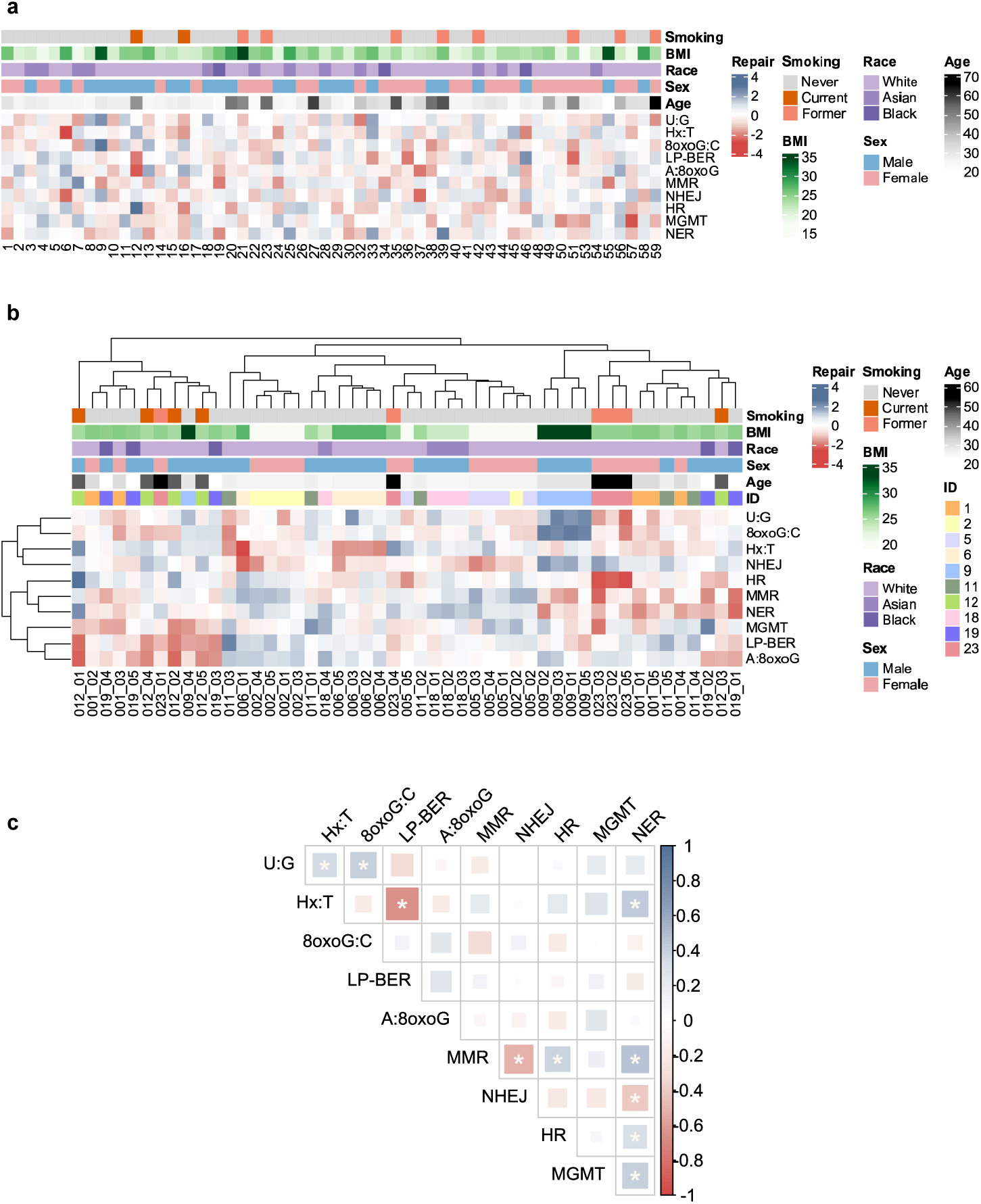
DNA repair capacity landscape. **(a)** DNA repair capacity landscape across 56 individuals. Standardized repair capacity was batch-corrected and averaged across replicates for each individual at their initial visit. Each column represents an individual and is labeled with their randomly assigned participant ID number. Demographics and other variables are indicated in the top six rows with colors corresponding to the key at right. The remaining rows represent Z-scored repair capacity as measured with the 10 FM-HCR reporter plasmids detailed in Table 1. **(b)** Hierarchical clustering of repeated measures in 10 individuals with multiple blood draws. Repair capacity was batch-corrected and averaged across replicates for each visit. The first three digits before the underscore symbol in the label correspond to the participant ID number; the last two numbers indicate the number of the blood draw. Participant ID numbers are also color-coded (see key at right) and reported in the row labeled “ID” **(c)** Pairwise Pearson correlation analysis of DNA repair pathways in 56 individuals at their initial visit. Multiple testing was corrected using the false discovery rate (FDR) with a significance threshold of p < 0.05.

**Fig. 4.**
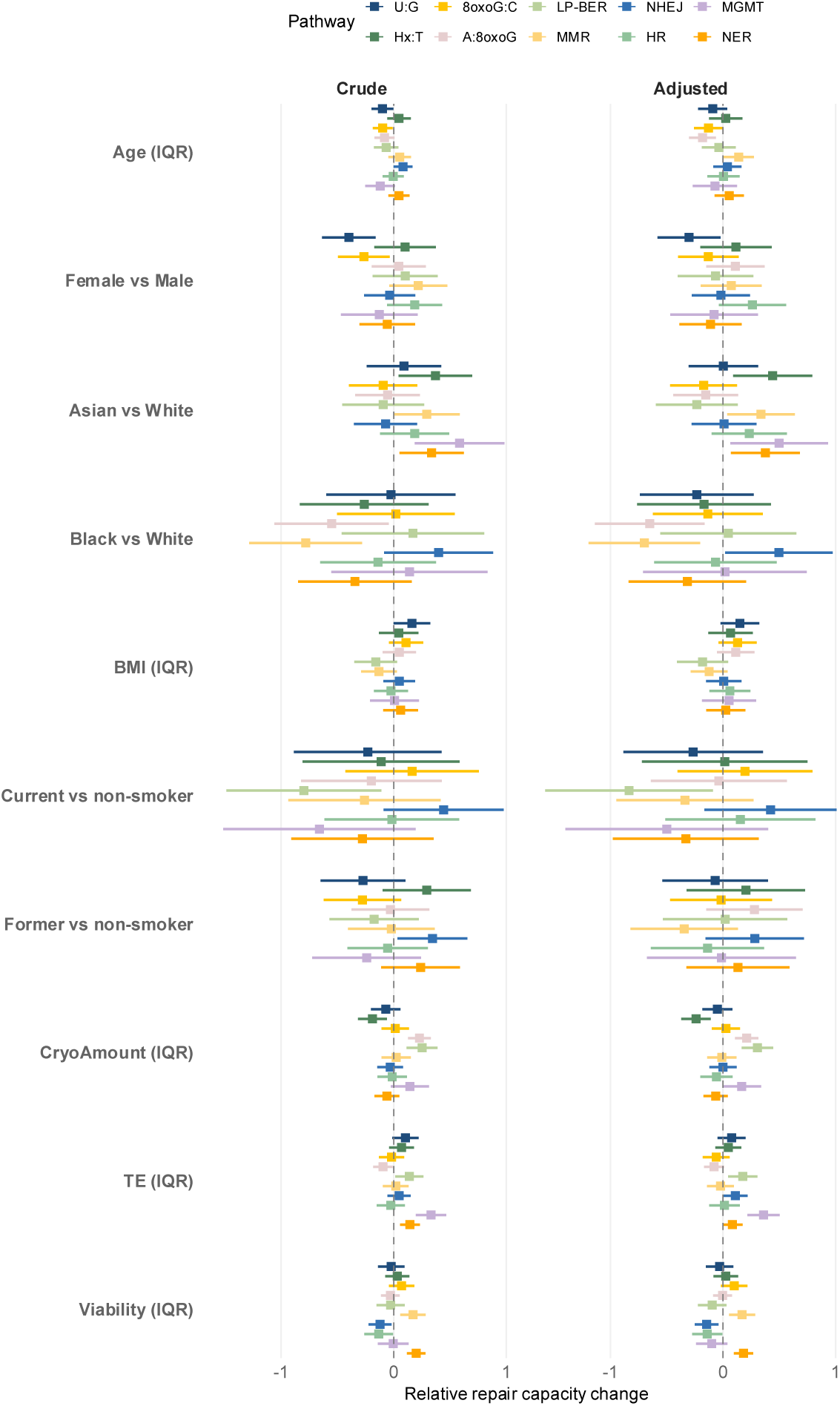
Associations between demographic or biological factors and DNA repair capacity. This plot represents the relative change in repair capacity for individual reporter assays (see key at top) associated with an increment of change in the value of other variables reported along the left side of the plot. A linear mixed-effects model was used to estimate the magnitude of change in z-scored repair capacity (expressed in standard deviations and reported on the horizonal axis) associated with changes in each examined variable. “Crude” refers to associations for unadjusted data; “Adjusted” refers to associations calculated after adjusting for covariates. For continuous variables, the boxes represent the change in repair capacity (expressed in standard deviations on the horizonal axis) corresponding to an interquartile range (IQR) increase in the variable at left. For categorical variables, the boxes reflect differences between the categories indicated at left. The lines on either side of the boxes represent the 95% confidence interval (95% CI). A 95% CI that does not overlap with the dotted vertical line at zero indicates statistical significance at the p < 0.05 level.

**Table 2.** Individual differences in DNA repair capacity.

| Pathway | Pop CV% | Sub-pop CV% | Ind CV% | P-value |
| --- | --- | --- | --- | --- |
| U:G | 21.8 | 22.5 | 14.8 | 2E-11 |
| Hx:T | 15.6 | 20.3 | 9.9 | 8.1E-16 |
| 8oxoG:C | 9.5 | 11.0 | 7.4 | 5.07E-11 |
| LP-BER | 11.7 | 12.4 | 8.2 | 1.48E-12 |
| A:8oxoG | 38.1 | 59.6 | 23.8 | 5.04E-28 |
| MMR | 15.1 | 17.5 | 10.1 | 1.49E-11 |
| NHEJ | 8.8 | 10.7 | 6.0 | 9.63E-12 |
| HR | 33.9 | 44.3 | 28.9 | 1.17E-06 |
| MGMT | 79.8 | 48.0 | 47.9 | 8.42E-09 |
| NER | 21.5 | 19.2 | 10.2 | 8.1E-16 |
| Average | 25.6 | 26.6 | 16.7 | - |

Note: Pop CV%: coefficient of variation % for inter-individual variation, calculated across 56 individuals' first visit. Sub-pop CV%: coefficient of variation % for inter-individual variation, calculated across the 10 individuals' first visit who had repeated visits. Ind CV%: coefficient of variation % for within-individual variation, calculated in participants with repeated visits. P value was from likelihood ratio test for inter-individual differences by comparing the linear mixed effect model with random individual component to the null model, after adjustment for multiple testing by false discovery rate (FDR).

Using a repeated measures study design, we evaluated the stability of DRC from multiple blood draws within same individuals over 4-6 weeks and observed clustering for most same-individual measurements (**Fig. 3b; Supplementary** Fig. 5a). The average across-pathway Euclidean distance within the same individual is 1.36 (**Supplementary Table 2**), while the average across-pathway distance among all individuals is 1.84 (**Supplementary Table 4**), further supporting clustering of same-individual measurements noted in **Fig. 3b**. Variability across the 10 individuals with repeated measures (average CV: 26.6%) was similar to variability across all 56 individuals (average CV: 25.6%) and larger than variability within the same individuals who had repeated measures (average CV: 16.7%) for all reporter assays (**Table 2**).

This highlights that DRC is relatively stable within individual over a short period while inter-individual variations are much larger.

### Co-regulation and competition among DNA repair pathways

Some correlations among DRC (**Fig. 3c**) were observed in data collected from the first visit for the 56 participants. Positive correlations between DRC measured by the U:G and Hx:T reporter assays (Pearson R = 0.35, FDR = 0.033) and the U:G and 8oxoG:C reporter assays (Pearson R = 0.42, FDR = 0.007), all three of which measure the initiation step of BER, suggest possible co-regulation of DNA glycosylases. LP-BER of the abasic site analog THF was inversely correlated with BER initiation measured by the U:G reporter assay (Pearson R = −0.30, FDR = 0.068) and the Hx:T reporter assay (Pearson R = −0.66, FDR = 3.5E-07), suggesting long-patch BER is less active in stimulated T-lymphocytes with higher glycosylase activities. NER was correlated with several other pathway activities (Hx:T, MMR, HR, MGMT), but was inversely correlated with NHEJ. Finally, MMR was positively correlated with HR (Pearson R = 0.39, FDR = 0.010) and NER (Pearson R = 0.47, FDR = 1.8E-03), and inversely correlated with NHEJ (Pearson R = −0.52, FDR = 3.3E-04), consistent with cell cycle regulation favoring MMR and HR in S-phase and G2M, while NHEJ is favored in G1 phase. It is possible that MMR, NER, HR, and NHEJ are co-regulated and the inverse relationship between NHEJ with the other pathways suggest competition for the same substrate.

To increase the robustness of our analysis, we tested whether the same correlations were reproduced when considering repair capacity data from repeated measures on an individual basis. Though the majority of associations tested in this manner were non-significant, positive correlations were observed between U:G with Hx:T and 8oxoG:C for 7 out of 10 individuals, LP-BER and A:8oxoG (9/10 individuals), NER with U:G (8/10 individuals), MMR (7/10 individuals), and HR (7/10 individuals); as well as MMR with HR (7/10 individuals) (**Supplementary** Fig. 5b). Inverse correlations were observed between LP-BER and Hx:T (10/10 individuals), LP-BER and U:G (8/10 individuals), NER and NHEJ (8/10 individuals), MMR and NHEJ (9/10 individuals). The associations in this separate set of data further supported the observed correlations among the 56 individuals reported in **Fig. 3c**. In a smaller dataset where the same reporter assays were performed in B-lymphoblastoid cell lines, only two associations reached statistical significance, but the direction of association was the same for 9/10 comparisons ^29^. In all cases, however, the most consistent observation is that the associations between different DNA repair activities are relatively weak and thus the activity of one DNA repair pathway cannot reliably be predicted from another.

### Relationship between DRC measured by CometChip and FM-HCR

Previously reported CometChip data for the repair of hydrogen peroxide (H_2_O_2_) induced DNA damage were re-analyzed to evaluate the effectiveness of batch correction and the Bayesian model fitting algorithm using within-individual CV of half-life estimates for the 10 participants with repeated measures. Compared to the traditional NLS method, the Bayesian algorithm reduced variation in repeated measures from 49.5% to 33.2% in data before batch correction, and from 28.6% to 23.1% in data after batch correction, suggesting a combination of batch correction and the Bayesian algorithm can improve the accuracy and robustness of DRC estimates (**Supplementary Table 5**). Similar to findings with FM-HCR, for CometChip data we observed greater inter-individual variability in CometChip data as compared to variability in measurements within the same individual (**Supplementary Table 6**).

**Fig. 5a** presents DNA repair kinetics at the first visit of the 56 participants, ranked by repair rate as indicated by half-life, and corroborated with corresponding demographics and DRC. Repair kinetics demonstrated greater heterogeneity among younger participants, with very fast or very slow repair kinetics observed more frequently (**Fig. 5b**). Among 12 participants aged 40 years and older, only 4 had slow repair kinetics, and none were classified as with very slow repair. Although a significant association was found between age and half-life (p = 0.0285), the observed patterns suggest a non-linear relationship with an uneven distribution of repair kinetics across age groups. There were trends suggesting differential kinetics by race and smoking status but the associations did not reach statistical significance, potentially due to inadequate sample size.

**Fig. 5.**
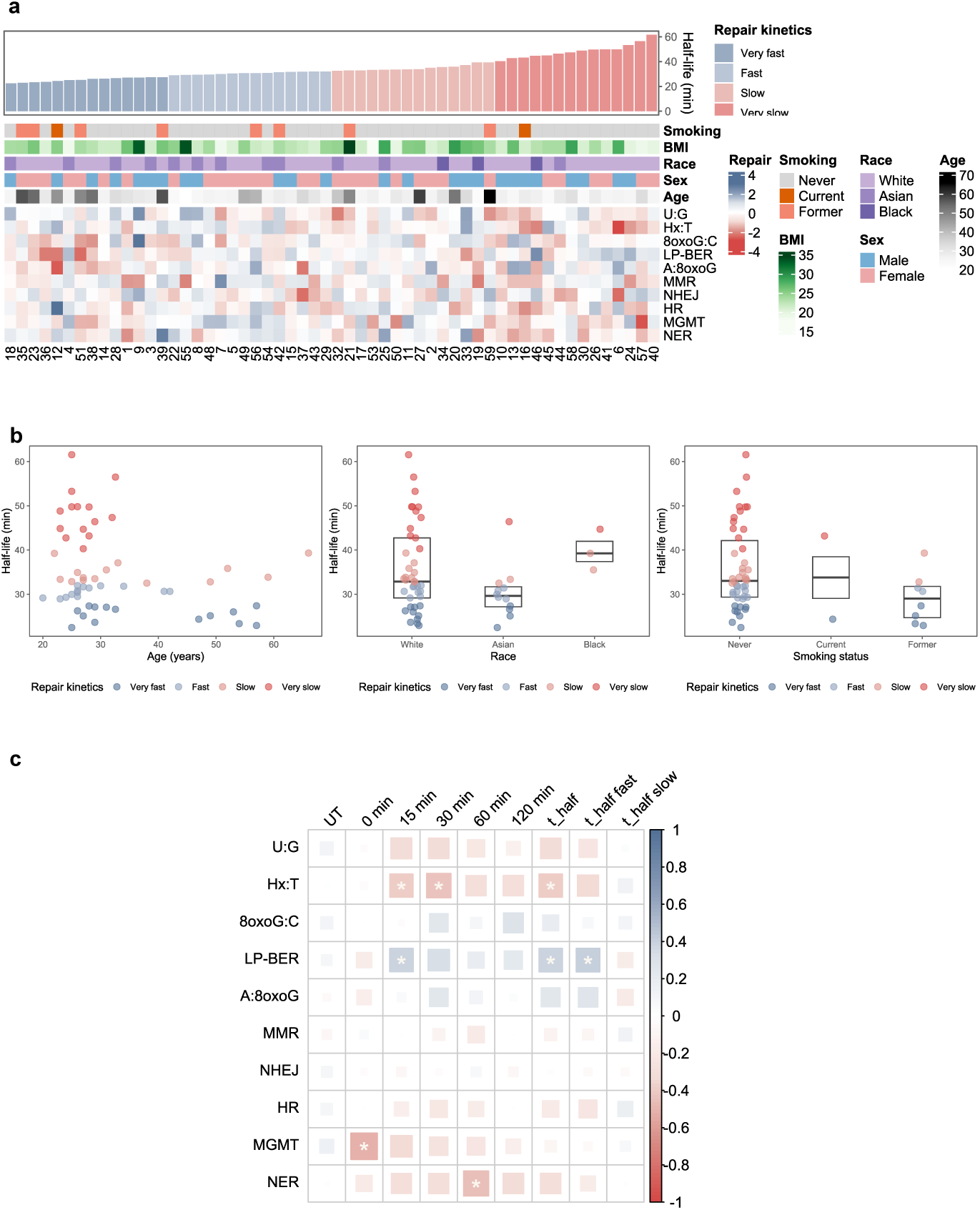
Visualization of DNA repair kinetics. **(a)** Heat map representing the relationships among DNA repair kinetics of PBMCs following treatment with H2O2 and other parameters. Each column represents data for PBMCs isolated from one of 56 individuals at their first visit and is labeled with their randomly assigned participant ID number. Demographics and other variables are reported in the top six rows with colors corresponding to the key at right. The remaining rows represent Z-scored repair capacity as measured with the 10 FM-HCR reporter plasmids detailed in Table 1. Individuals were ordered and grouped by half-life (t1/2) estimates (Equation S0), from the smallest (fastest repair) to the largest (slowest repair). **(b)** Scatter plots showing borderline significant associations between t1/2 estimates and demographic variables among the 56 individuals. **(c)** Spearman’s correlation analysis between DNA damage levels between 0 and 120 minutes after treatment of PBMCs with H2O2 and repair kinetics parameters obtained by fitting CometChip data to Equation S0. Spearman’s R is represented according to the color scale at the right of the plot, and larger boxes represent larger absolute values. Statistically significant associations are marked with a white asterisk. Multiple testing was corrected using the false discovery rate (FDR) at a significance level of 0.05.

Correlations between DRC with damage levels and repair kinetics are presented in **Fig. 5c**. MGMT was found to be inversely correlated with initial damage levels after H_2_O_2_ treatment (Spearman R = −0.50 at 0 min, FDR = 0.008) while Hx:T and NER were inversely correlated with damage levels at later time points (Spearman R = −0.42 for Hx:T at 30 min, FDR=0.034; and −0.45 for NER at 60 min, FDR = 0.020) and LP-BER was positively correlated (Spearman R = 0.38 at 15 min, FDR = 0.050) with damage levels. There was an inverse correlation between the time required to repair 50% of the induced DNA damage (t_1/2_) for Hx:T (Spearman R = −0.39, FDR = 0.039) indicating that repair kinetics are faster for individuals with more efficient repair of the Hx:T lesion. By contrast, there were positive correlations between both overall (t_1/2_) and fast (t_1/2_ fast) phase repair kinetics and LP-BER (Spearman R = 0.41, FDR = 0.034 for both half-life estimates), indicating an inverse relationship between the rate of repair for H_2_O_2_ induced DNA damage and the efficiency of repair for the THF lesion. To further evaluate these correlations at the individual level, we examined repeated measures from the 10 participants. When comparing correlations between repair processes in the full cohort of 56 individuals to correlations between repair processes within the subset of 10 individuals with repeated measures, some patterns emerged. Notably, 8 out 10 individuals showed a correlation between *higher* LP-BER with *higher* damage levels at 30 min and 60 min, as well as slower repair of H_2_O_2_ induced damage as reflected in *longer* half-life estimates over repeated measures (non-significant) (**Supplementary** Fig. 5c).

## Discussion

Functional assays measuring the entire DNA repair landscape present an opportunity to gain profound insights into human variation in genome maintenance, but they must first be rigorously tested and validated. Here we report a comprehensive analysis of DRC in six major pathways in PHA stimulated PBMCs. A repeated measures study design and systematic consideration of both biological and technical variables establish FM-HCR as a powerful approach for measuring DRC in populations. Variation between individuals can be explained in part by lifestyle and demographic variables and is larger than within-individual variation, consistent with findings others have made using alternative assays. Our findings furthermore establish that the activity each DNA repair pathway is largely independent of others, reinforcing previous observations in immortalized cell lines and underscoring the importance of analyzing multiple pathways for a complete picture of genome integrity ^29,30^. We find, as expected, that comet assays provide complementary information about DNA repair in primary lymphocytes, and variation in the ability to repair H_2_O_2_ induced genomic DNA damage can be explained in part by differences in DRC measured by FM-HCR. In addition to these biological findings, we report useful code and analytical frameworks that support population DNA repair studies with both comet assays and FM-HCR. Here we discuss in more detail the technical considerations and biological findings that are important for future studies.

### Within-individual stability and inter-individual variation

Our study design included repeated measures that allowed us to assess whether DRC as measured by FM-HCR is sufficiently stable over time to draw meaningful conclusions about inter-individual variation. Indeed, we observed significant inter-individual variability across all DRC assays, contrasting with relative within-individual stability. The 38.1% population CV observed in MUTYH (A:8oxoG) repair activity is striking given MUTYH’s critical role in responding to oxidative DNA damage ^21,33^, as well as the increased risk for MUTYH-associated polyposis (MAP) even among individuals carrying a single inactive allele ^10,34–38^. The observed inter-individual differences in MUTYH activity reflect a spectrum of repair efficiencies that could be influenced by genetic variants, gene expression, or post-translational modifications ^39^. To our knowledge, aside from a previous study on MUTYH-associated variants ^21^, ours is among the first to report functional variation of MUTYH in PBMCs from a healthy cohort. These findings warrant further investigation to determine whether such variation contributes to differential cancer susceptibility, particularly in individuals with monoallelic or biallelic MUTYH mutations.

The small inter-individual differences observed in NHEJ (population CV = 8.8%) could be an indirect consequence of relatively fast repair by this pathway ^40^. We note that the timepoint selected for our assays (24 hours) occurs long after repair via this pathway is expected to be complete; future studies including earlier time points might better resolve small differences in the activity of NHEJ between individuals. In contrast, the higher variability in HR (population CV = 33.9%), which is much slower than NHEJ, and may reflect biological variation in cell cycle dependent regulation: NHEJ is used almost exclusively in quiescent cells, whereas HR is active during cell proliferation ^41,42^. These findings suggest a need for future studies investigating repair kinetics.

Immortalized lymphoblastoid cell lines (LCLs) have been used as models for measuring inter-individual variation in DRC, but the extent to which they recapitulate the variability in primary cells has been unclear. We previously used FM-HCR in a panel of 24 Epstein-Barr virus transformed B lymphoblastoid cell lines. Comparing our previous data with new data in PHA-stimulated PBMCs, we observe that some pathways like A:8oxoG repair and NER show comparable variability in both primary cells and LCLs ^29^, whereas others, particularly MGMT activity, differ significantly. MGMT activity was uniformly high in primary lymphocytes with % reporter expression generally less than 0.1%, comparable to a previously reported positive control, TK6+MGMT ^43^. By contrast, MGMT activity was highly variable in LCLs ^9^; indeed for some LCLs MGMT activity was less than 5% of the level observed in a TK6+MGMT positive control ^43^. MGMT silencing in cancer has long been known ^44^, but these findings suggest it can occur in immortalized cells outside the context of carcinogenesis. Our data underscore the importance of validating findings regarding human variation from immortalized cells in primary cells. To make more direct comparisons with previous work, future studies would benefit from including other immune cell types (e.g., B cells, NK cells, and monocytes) and comparing both proliferating and quiescent cells.

Compared to existing studies, our findings on inter-individual variability were generally lower (**Supplementary Table 8**). Specifically, variability in Hx:T repair activity (predominantly initiation of BER by the AAG DNA glycosylase) ^45–47^, NHEJ ^48^, OGG1 ^49,50^, APE1 ^51,52^, NER ^7^, and UNG ^53^, was lower in our study. In contrast, the high variability in MGMT repair activity aligned with high variability reported previously across quiescent cell free extracts ^54^ and human organs ^53^. Differences in the extent of variation observed by FM-HCR in our study population versus previous reports may be explained by a combination of different assay techniques, differing population demographics, the use of PHA stimulated PBMCs, and the potential variability in cell free extracts due to multiple cell types that vary in abundance across populations and have different DNA repair capacities ^55^. These further highlight the need to establish both a standardized sample type and handling strategy that reliably reflect individual DRC.

The high CV observed for MGMT in our dataset may be partly influenced by zero signals in some assays due to extremely low reporter expression (approximately 0.00749% in raw data). Since expression of this reporter is inversely proportional to MGMT activity ^30,43^, the absence of signal can be interpreted to mean the activity of lymphocytes was too high to be measured with the amount of reporter plasmid used in our assays. In a subset of individuals, we performed assays with a large excess of the MGMT reporter plasmid (2 μg), which resulted in a more robust signal (10-fold increase) that correlated (Pearson R = 0.84, p = 0.009) with findings using smaller amounts of plasmid (500 ng) (**Supplementary** Fig. 6a), but this analysis was not feasible to carry out in the larger population. Including zero values without correction can bias estimates by artificially inflating within-person variation relative to inter-individual variation, thus obscuring true biological differences. Future studies may therefore consider implementing minimum detection thresholds or alternative assay methods that produce a more robust signal in cells with high levels of MGMT activity to mitigate this issue.

### Associations between demographic variables and DRC

Previous studies indicate DRC is influenced by the environment, genetics, and age. We utilized linear mixed-effects models to investigate associations between demographic variables and inter-individual variability. Our analysis identified a significant inverse relationship between BER capacity and participants’ age, particularly A:8oxoG repair capacity, while an association with a decline in 8oxoG:C repair capacity remains borderline after adjusting for covariates. Previous work indicates DRC may decline with age, but in a manner that may depend on cell type, genetics, and other variables ^3,8^. Recent studies indicate both MUTYH-dependent A:8oxoG repair ^56^ and OGG1-dependent 8oxoG:C repair ^57^ affect telomere length, a well-established biomarker of aging that also shortens under oxidative stress ^58^, and is protected from oxidative stress by DNA repair machinery ^59^. Future work is needed to determine whether BER capacity associates with telomere length in populations. Although prior work has reported age-related declines in NER and MMR in cells and tissues ^8^, we did not observe such reductions in our cohort. Since our study population is relatively young, with an average age of 28 years and no participants over the age of 60, some age-dependent changes may not be observable in the population we studied ^60^.

Although our sample size was small and not well-powered to investigate associations between race and DRC, some preliminary associations were observed across several DNA repair pathways, including BER pathways (Hx:T, A:8oxoG), MMR, and NER. Previous literature on racial diversity has been focused on genomic variation across racial groups ^61,62^, and there are still relatively few studies that include functional measurements that reflect environmental drivers of health disparities. Our primary findings should be interpreted cautiously, as they may reflect underlying social or environmental factors and require validation in larger, more diverse cohorts. We also did not replicate a previously reported association between reduced NER activity with increasing BMI ^63^, likely due to the limited number of participants in our study with a BMI >25 kg/m^2^, a threshold beyond which BMI-related effects are typically evident ^64^. Additionally, smokers in our cohort exhibited lower repair of our LP-BER reporter, with a substantial effect size (est. = −0.83), though no longer significant after multiple-testing correction (FDR = 0.258). This trend is consistent with prior reports linking smoking and oxidative stress to diminished DNA repair ^6566,67^. Overall, our data add to a large body of evidence indicating that lifestyle and environmental factors can affect genome integrity mechanisms.

### Correlation among DNA repair pathways

While there were some correlations between DNA repair pathways, a lack of strong correlations between DNA repair pathways is the most notable trend in our data. FM-HCR reporter assays have previously been validated rigorously in repair deficient cell lines. This validation has repeatedly confirmed that each assay reports DRC for a specific pathway and in a manner that is independent of defects in other pathways, except when two or more pathways compete for the same DNA lesion (e.g. HR and NHEJ competing for repair of a DSB). Although we cannot completely rule out technical sources of bias, correlations between reporter assays may reasonably be assumed to reflect co-regulation of repair activities. While most of the correlations we observed among reporter assays in PBMCs (n = 56) did not reach significance in lymphoblastoid cell lines (n = 24), most trends were recapitulated ^29,30^. Specifically, lymphoblastoid cell lines yielded positive associations between U:G and Hx:T, U:G and 8oxoG:C; NER and MMR, NER and HR; MMR and HR, and negative associations between LP-BER and U:G, LP-BER and Hx:T; and MMR and NHEJ.

Overall, the lack of strong correlations across all pathways highlights the complexity and specialization of DNA repair mechanisms, and underscores the importance of monitoring each pathway independently. This independent assessment provides rich information on pathway-specific functions, inter-pathway crosstalk, and the regulatory networks governing DNA repair. By leveraging a variety of reporter plasmids tailored to specific pathways through our FM-HCR assays, we can comprehensively analyze repair capacity, identify deficiencies, and uncover dysregulation that may underly disease states linked to genomic instability.

### Associations between FM-HCR and DNA repair kinetics measured by comet assay

Although CometChip and FM-HCR are run on different time scales and with different types of DNA damage, we sought to identify associations between them that might help to reinforce the interpretation of our dataset. The level of genomic DNA damage measured by CometChip at t_0_ reflects the combined effects of BER initiation, which increases the number of strand breaks, and repair completion, which decreases the number of strand breaks. Inefficient processing of oxidized base lesions and abasic sites can result in persistent DNA damage due to the accumulation of BER intermediates, whereas cells with more efficient BER systems resolve these intermediates before t_0_. The repair of oxidized bases induced by H_2_O_2_ occurs mainly via LP-BER ^68^, but we observed an association between LP-BER capacity with *higher* damage levels and *delayed* repair, suggesting that increased LP-BER activity leads to slower overall repair of H_2_O_2_-induced DNA damage. H_2_O_2_ induces a spectrum of DNA lesions, including single-strand breaks, but the majority are oxidative base lesions ^69^.

Short-patch BER is a relatively rapid process, with repair typically completed within approximately 20 minutes in cells ^68^, whereas long patch BER is slower. Collectively, our data are consistent with a model in which long patch BER competes with other faster pathways for the repair of peroxide-induced damage. In such a model, the overall rate of repair would be inversely proportional to the amount of flux through the LP-BER pathway.

The inverse relationship we observed between t_1/2_ and NER is consistent with its reported role in repairing some types of H_2_O_2_-induced DNA damage ^69,70^. Although the alkyl lesions repaired by the glycosylase that repairs Hx:T are not major products of oxidative damage induced by H_2_O_2_, we observed significant inverse correlations between Hx:T repair and both DNA damage levels and repair rates. This apparent association between AAG activity (as measured by the Hx:T assay) and the resolution of H_2_O_2_-induced DNA damage could reflect repair of etheno adducts that occur following lipid peroxidation ^71^. Etheno adducts are induced by H_2_O_2_ in lymphocytes ^72^, and are repaired by both AAG and NER ^73^. Additionally, MGMT activity inversely correlated with initial levels of H_2_O_2_-induced DNA damage. Since MGMT does not directly repair H_2_O_2_-induced damage, this correlation suggests an indirect link. Notably, several studies report an association between levels of MGMT and the antioxidant glutathione ^74–76^, which protects against peroxide-induced DNA damage. MGMT levels might therefore be at least a partial surrogate for glutathione levels in PBMCs, and this would be consistent with the inverse relationship between MGMT activity and initial levels of peroxide-induced damage.

### Important limitations

We acknowledge several limitations in our study. First, our analysis focused on PHA-stimulated lymphocytes, which are expected to exhibit different levels of variability and DNA repair capacities compared to resting lymphocytes due to proliferation.

Lymphocytes also may not be representative of cells from other tissue types. Future research should explore repair capacities between stimulated and resting lymphocytes and extend analyses to multiple cell and tissue types to address this question. If it can be established that DNA repair in PBMCs is a reliable proxy for genome maintenance in other cell types, these assays could have major applications in personalized treatment and prevention of diseases that are caused by genome instability. Second, although we established a comprehensive data processing pipeline and successfully addressed technical variation through batch effect correction and linear mixed-effect models, some variability related to batch differences may still persist. To further minimize technical noise, future studies should aim to maximize the number of samples included in each batch during the study design phase, coupled with rigorous sample quality control measures. Third, our cohort predominantly comprised younger individuals with high socioeconomic status, limiting the generalizability of our results to broader populations. As we discovered that DRC could vary with demographic factors, future studies should include a more diverse and representative sample to ensure the findings are applicable to the general population. Fourth, the study monitored within-person variation over several months, potentially missing long-term or age-related changes in DNA repair mechanisms. Longitudinal studies with extended follow-up periods are necessary to capture temporal variations and the impact of aging on DRC. Lastly, aside from the CometChip assay, we did not incorporate multi-modal data to thoroughly investigate the complex regulation of DNA repair mechanisms, such as genetic, transcriptomic, and proteomic factors and phenotypes including environmental exposures and health outcomes. Future studies should integrate these data types to address these gaps and achieve a more comprehensive understanding of inter-individual variation in DRC to better aid precision medicine.

### Novelty and significance

Our findings have several important implications for the field of DNA repair and precision medicine. The standardized and validated assays in this study can serve as sensitive benchmarks for DRC measurements in human-based studies. By demonstrating the reproducible detection of subtle inter-individual differences, our study establishes the feasibility of integrating FM-HCR into larger-scale molecular epidemiological studies, potentially uncovering previously unrecognized associations between DNA repair, disease risk, and health outcomes. Furthermore, we established a comprehensive data processing and standardization framework to control technical variation and ensure consistency across future DRC studies. To facilitate broader adoption, we developed a user-friendly web interface that allows researchers without a computational background to analyze and interpret DRC data, bridging disciplinary gaps between laboratory-based researchers, epidemiologists, and computational scientists.

The ability to differentiate individuals based on DRC also paves the way for mechanistic studies exploring how genetic background, demographic factors, and environmental exposures influence DNA repair processes. These insights can inform the development of targeted interventions aimed at enhancing DNA repair mechanisms and maintaining genomic integrity, and predicting the health effects of exposure to DNA damaging agents based on individual repair profiles.

## Methods

### Study participants

The study enrolled 56 healthy volunteers at the Clinical Research Center at the Massachusetts Institute of Technology in Cambridge, MA. Of these volunteers, 10 participated in a repeated measures study involving the collection of 4 to 5 blood samples over a period of 4 to 6 weeks. The remaining 46 participants underwent a single blood draw. Demographic information was collected at the time of enrollment using a questionnaire that included age, sex, race, height, weight, smoking history, alcohol consumption, family history of cancer, place of birth. Participants were recruited using a flyer that was posted in campus buildings and offered $25 for the first three visits and $35 for subsequent visits. The study was approved by the Committee on the Use of Humans as Experimental Subjects at MIT (COUHES protocol #1306005796), and all participants provided informed consent prior to participation.

### Sample processing

We have described blood sample processing, cryopreservation, recovery and culture for this study in our previous publications ^29,32^. Briefly, peripheral blood mononuclear cells (PBMCs) were obtained from the buffy coat of fresh whole blood using standard Ficoll gradient density centrifugation (GE Healthcare Bio-Sciences). PBMCs from each participant were cryopreserved in 1 mL aliquots with at least 5×10^6^ cells per vial in freezing medium composed of 40% RPMI-1640 (ThermoFisher Scientific, Waltham, MA), 50% heat-inactivated fetal bovine serum (FBS; Atlanta Biologicals, Inc., Flowery Branch, GA), and 10% dimethyl sulfoxide (DMSO; MilliporeSigma, St. Louis, MO). Cell stocks were initially cooled to-80 °C at a rate of approximately 1 °C per minute, and transferred to liquid nitrogen storage 24 hours later. Four separate vials from each blood sample were stored for future experiments, with three vials designated as replicates for reproducibility assessments.

Cryopreserved PBMCs were recovered following standard procedure as described previously, yielding cell viability rates exceeding 85%, as determined by an automated Trypan Blue exclusion system (Vi-CELL cell counter). T-lymphocytes were then stimulated with Phytohemagglutinin-L (PHA-L; MilliporeSigma, St. Louis, MO) in stimulation media (RPMI 1640 + 20 % FBS + 1% penicillin/streptomycin + 5µg/mL PHA-L) for 3 days prior to conducting the FM-HCR assays.

### Study workflow and experimental design

The primary objectives of this study were to evaluate the sensitivity of the FM-HCR assay in detecting inter-individual variation in DRC and to assess the within-individual stability of DRC over time. Accordingly, our experiments were designed to evaluate and distinguish biological variability from technical variation arising from sample processing and experimental conditions. FM-HCR assays were conducted across 37 batches over 57 days (**Fig. 1**). Each batch comprised a distinct subset of individuals to facilitate the assessment of inter-individual variation while minimizing batch-related confounding. For participants with multiple blood draws, we evaluated the within-individual stability of DRC by analyzing repeated measures for each participant. Key technical details and sample-specific variables recorded for each sample included:

(1) batch: an identifier for each FM-HCR assay batch; (2) CryoTime: the duration of storage in liquid nitrogen (in days); (3) CryoAmount: the number of cells available for cryopreservation after isolation; (4) cell viability: the percentage of live cells following transfection; (5) transfection efficiency (TE): the percentages of cells that expressed a transfection control plasmid in FM-HCR assays. The effect of each variable was determined through a combination of prior knowledge on their impacts and observed associations, as detailed in **Supplementary methods**. For each participant’s blood draw, three replicate samples were distributed across different batches to evaluate assay reproducibility and the impact of batch variability on measurements. Additionally, replicate samples from fresh whole blood samples by an anonymous healthy donor (Research Blood Components, Brighton, MA) were included in each batch as internal controls, providing additional data points for assessing data consistency across batches.

### FM-HCR assays

DRC was measured by FM-HCR with 10 reporter plasmids targeting specific DNA repair pathways (**Supplementary Table 9**). The detailed methodology of the FM-HCR assay has been described previously ^29,30^. Briefly, the FM-HCR assay quantifies the ability of cells to repair specific types of DNA damage introduced into reporter plasmids, with repair efficiency reflected by the expression levels of fluorescent reporter genes. Our assays targeted pathways including: MUTYH-dependent initiation of base excision repair (BER) of a normal adenine opposite an 8oxoG lesion (A:8oxoG); OGG1-(and likely NEIL1-and NEIL2-) dependent BER initiation of a site-specific 8oxoG lesion opposite cytosine (8oxoG:C); a UNG-(and likely SMUG and TDG) dependent BER of a uracil lesion opposite guanine (U:G); AAG-dependent initiation of BER for a hypoxanthine DNA lesion opposite thymine (Hx:T); long patch base excision repair (LP-BER) of an abasic site analog tetrahydrofuran; nucleotide excision repair (NER) of UV-C light-induced DNA damage; non-homologous end joining (NHEJ)-dependent repair of a double-strand break; mismatch repair (MMR) of G:G mismatch; methylguanine methyltransferase (MGMT)-dependent direct reversal of a *O*^6^-MeG:C lesion; homologous recombination (HR)-dependent repair of a blunt-end double-strand break. Throughout this manuscript, each assay is hereafter referred to using the abbreviations defined in this paragraph.

Stimulated PBMCs were combined with plasmid cocktails and transfected via electroporation using a 96-well Bio-Rad MXcell Gene Pulser with an exponential waveform at 260 V and 950 μF. Each well contained 2 × 10^6^ cells in 100µL complete medium (RPMI 1640 + 20% FBS + 1% penicillin/streptomycin). Transfected cells were incubated at 37 °C with 5% CO_2_ for 24h prior to flow cytometry. Flow cytometry was conducted on a BD LSR II cytometer as previously described ^29,30^. To establish gating for positive cell populations, both single-color controls and dropout controls (excluding the reporter of interest during transfection) were utilized. If a flow cytometry run yielded fewer than 30 fluorescent events, the experiment was repeated to ensure data reliability. We then took the percentage ratio of reporter expression from damaged reporter plasmids to that of corresponding undamaged reporter plasmids as detailed in our previous publications. These FM-HCR percent reporter expression values were used as the raw data input for the subsequent data pre-processing pipeline for controlling for technical variability.

### Data pre-processing

We developed a standardized data processing pipeline for the application of FM-HCR in population studies (**Fig. 1**). To mitigate the impacts of outliers and achieve normalization, the raw reporter expression data was first natural-log transformed ^77^. For five reporter assays (i.e., U:G, Hx:T, 8oxoG:C, A:8oxoG, MGMT), higher reporter expression corresponds to lower DRC because the fluorescent signal arises from transcriptional errors induced by the unrepaired DNA lesion ^29,30^ For these assays we therefore performed sign inversion after log-transformation so that higher transformed values consistently corresponded to higher DRC across all pathways. To unify the scale and facilitate cross-pathway comparisons, the data were further standardized by z-score conversion. The raw reporter expression of the MGMT pathway was on average below 1%, and some measurements yielded a zero reading, indicating that signal was below the detection limit. These data were considered as missing values with a missing rate at 18.2% and imputed from the average of other replicates from the same individual to preserve individual-level data integrity during batch correction.

Potential batch effects were evaluated using strategies tailored to our study design (**Supplementary methods**). Batch correction was performed using the ComBat function from the ‘*sva*’ R package ^78^, employing parametric adjustments based on the batch indicator while preserving inter-individual variability by accounting for age, sex, race, BMI, and smoking status. The effectiveness of batch correction was evaluated using variation metrics, including coefficients of variation (CV) across batches and within individuals.

### Comet kinetics modeling

The CometChip assays were performed in parallel with FM-HCR following the same experimental design and have been previously described in detail ^32^. In this study, we revisited the DNA repair kinetics data after applying batch correction outlined in the previous section. We also enhanced the kinetics model fitting by employing a Bayesian inference approach to estimate the posterior distributions of the model parameters, which reduces the impact of unstable estimates that can arise from traditional non-linear least squares (NLS) algorithms. Specifically, background-corrected DNA damage levels were modeled using a biphasic exponential decay function, which accounts for both fast and slow phases of repair. The model is formulated below:

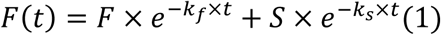

Where F(t) is damage level at time t, F and S are initial damage levels and k_f_ and k_s_ are rate constants for the fast and slow phases of repair kinetics. All parameters were constrained to be larger than 0 and assigned weakly informative normal priors based on our empirical knowledge of how hydrogen peroxide-induced DNA repair decay over time (**Supplementary methods**). Posterior sampling was performed using four Markov Chain Monte Carlo (MCMC) chains (2000 iterations each; 1000 warm-up) in the ‘*brms*’ R package ^79^. Convergence was confirmed by examining Gelman-Rubin statistics (all < 1.01), and model performance was evaluated using Leave-One-Out Information Criterion (LOOIC). We then estimated t _1/2_ (half-life), the time required to repair half of initial damage, through numerically solving the biphasic decay equation for each sample.

## Statistical analysis

Two participants (3.6%) had missing age data, and four participants (7.1%) were missing height or weight measurements necessary for calculating body mass index (BMI). To maintain statistical power and minimize bias, we performed multiple imputation under the assumption of data being “missing at random” (MAR). Missing values were imputed using the Multivariate Imputation by Chained Equations (MICE) method, incorporating relevant demographic and biological variables into the imputation model, including age, sex, race, BMI, smoking status, CryoAmount, cell viability, and TE. Twenty imputed datasets were generated, and effect estimates from the subsequent regression analyses were pooled using Rubin’s rules to account for imputation uncertainty ^80^. Additionally, a sensitivity analysis was conducted to compare the results of the complete case analysis with those obtained from the multiple imputed datasets.

We employed linear mixed-effects (LME) models to detect inter-individual variation in DRC and explore contributing factors. Random intercepts for individuals were included to account for correlations in repeated measures. The general LME formula is:

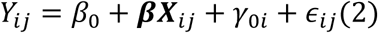

Where *Y_ij_* denotes the DRC measurement for individual i at visit j; β_0_ denotes the fixed inter-individual intercept; γ_0*i*_ denotes the random intercept; and ϵ*_ij_* denotes the random noise. ***β*** is a vector of coefficient(s) for covariate(s) ***X**_ii_*.

We used four models to explore associations: model 1 (null model) evaluated inter-individual variation by comparing to a model excluding γ_0*i*_; model 2 (univariate model) assessed crude associations between each variable ***X**_ij_* and DRC; model 3 (multivariate model – fixed effects) evaluated adjusted associations of demographics ***X**_i_*with DRC; and model 4 (multivariate model - mixed effects) evaluated adjusted associations of mixed technical-biological factors ***X**_ij_* and DRC, controlling for demographics ***X**_i_*. Demographics considered in models 3 and 4 included age, sex, race, BMI, and smoking status, selected based on significant crude associations with DRC. Detailed formulas for each model can be found in the **Supplementary methods**.

We evaluated the within-individual stability of repeated measures using hierarchical clustering and calculated the Euclidean distance across the 10 reporter assays to provide a numerical metric for clustering. Specifically, the averaged distance across repeated measures within the same individual was compared to the distance across individuals to evaluate the proximity of data points. A general formula for the distance between repeated measures a and b within the same individual or between averaged measures from individuals a and b is:

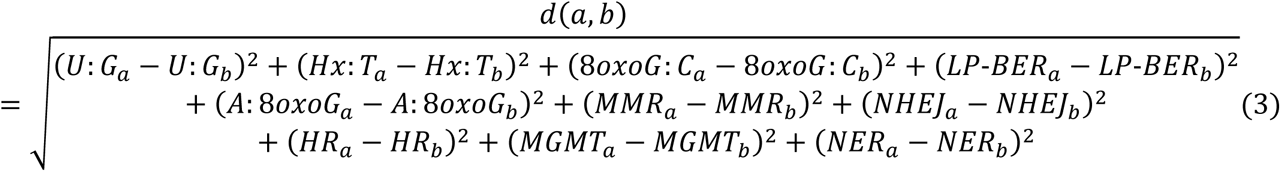

In this equation, *d(a,b)* represents the Euclidean distance, *DRC_a_* represents the value of the first measurement, and *DRC_b_* represents the value for second measurement of a particular reporter assay.

The correlation between DNA repair assays was explored by Pearson’s correlation and correlations between repair capacity and other factors was explored by Spearman’s correlation. Multiple testing correction was performed using false discovery rate (FDR) with a significance threshold of 0.1 ^81^, accounting for 45-90 correlation tests and 10 LME models, each assessing a different DRC outcome. All analyses were performed in R (version 4.3.2). The process of FM-HCR data standardization, batch effect correction, and comet kinetics modeling was incorporated in and performed through our R package ‘*pdrc’ [*https://github.com/NagelLabHub/pdrc].

## Data availability

The data supporting the findings of this study are available within the published article, supplementary information, or source data file.

## Code availability

R scripts for analyses and figures can be found at https://github.com/NagelLabHub/COUHES and the R package and website we developed can be accessed at https://github.com/NagelLabHub/pdrc.

## Acknowledgements

The authors acknowledge funding that supported this work from the National Institute of Environmental Health Sciences (U01ES029520 to Z.D.N. and DP1ES022576 to L.D.S.).

## Author contributions

Conceptualization: I.A.C., Z.D.N., L.D.S.

Investigation: I.A.C., Z.D.N., P.M., T.Z., C.R.

Methodology, analysis, visualization, software, writing – original draft: T.Z. Writing – review & editing: D.C.C, L.L., I.A.C., Z.D.N.

Supervision and funding acquisition: Z.D.N., L.D.S.

