## Supplementary for "Comprehensive Measurement of Inter-Individual Variation in DNA Repair Capacity in Healthy Individuals"

### Supplementary Methods

#### Strategies for batch effect evaluation

To ensure the reliability of our FM-HCR measurements and accurately assess inter-individual variations in DNA repair capacity (DRC), we developed a comprehensive strategy to evaluate batch effects and the efficiency of batch effect correction. The evaluation comprised the following steps:

1. Visual Inspection of Measurements Across Batches:

We visually inspected standardized DRC measurements across batches and individuals. In the absence of batch effects, standardized data should be centered around the population mean (standardized to zero) across all batches, with variability arising only from inter-individual differences and random noise. Replicates from the same individual should cluster closely together. Before batch correction, we observed shifts in measurements by batch (**Supplementary Fig. 1a**) and significant variations within the same individual (**Supplementary Fig. 2a**). After batch correction, data were re-centered batch-wise (**Supplementary Fig. 1b**) and within-individual variability was reduced (**Supplementary Fig. 2b**).

2. Evaluation of Variations Across Batches:

We calculated the coefficient of variation (CV) to quantify variability across batches and within individuals. Although it can be intuitive to report variability in terms of the fold-range of DRC, we employed the more robust metric CV to quantify variability. CV normalizes variability relative to the mean, enabling meaningful comparisons across biomarkers with different expression levels [1]. Moreover, CV is less sensitive to extreme values than fold-change, making our assessments more reliable and representative, particularly in large-scale studies where outliers can disproportionately influence fold-change measurements. CVs were computed on the original reporter expression scale of the data since standardized data with a mean of zero cannot be used for CV calculation. For across-batch CV, we calculated the standard deviation of batch means across batches divided by the average batch mean. For within-individual CV, we computed the standard deviation of samples divided by their mean for each individual, then averaged these values across all individuals. *Before Correction:* Across-batch CVs ranged from 12.4% to 45.7% (mean 25.8%) among the 10 pathways; within-sample CVs ranged from 15.9% to 46.5% (mean 28.6%) (**Supplementary Table 1**). *After Correction:* Across-batch CVs ranged from 2.7% to 18.2%, and within-sample CVs ranged from 9.0% to 46.7%. All pathways except MGMT achieved reductions in within-sample variability.

<u>3. Correlation Analysis Between Batch Indicators and Measurements:</u>

We evaluated the correlation between batch indicators and DRC measurements. A strong correlation indicates significant batch effects. *Before Correction:* We observed a strong correlation between batch indicators and DRC measurements (**Supplementary Fig. 3a**). *After Correction:* These correlations were substantially reduced or absent, demonstrating effective batch effect removal (**Supplementary Fig. 3b**).

<u>4. Assessment Using Internal Control Samples:</u>

We further evaluated batch correction efficiency using internal control samples - PBMCs from an anonymous healthy volunteer included in every batch. *Before Correction:* CVs ranged from 17.8% to 98.3% (mean 35.8%) (**Supplementary Table 1**). *After Correction:* By adjusting for sample types using the ComBat function, the CVs decreased to a range of 10.6% to 73.5%, with a mean reduction of 24.3%. We investigated whether including these internal control samples in the batch correction process for the experimental samples would enhance result stability. However, due to the anonymous nature of the control individual and the presence of only one replicate per batch, this approach introduced random noise that adversely affected the stability of the experimental samples (**Supplementary Table 1**). Consequently, we performed batch effect correction using only the experimental samples (experimental-sample-only) for our downstream analysis.

<u>5. Additional evidence in clustering of repeated measures:</u>

Although time-related variations may contribute to overall variability, clustering of repeated measures provides additional evidence for the efficacy of batch correction. We evaluated within-individual variability by comparing the CV and Euclidean distance of repeated measures before and after batch correction. The CV of repeated measures reduced from 20.1% to 16.7% and the distance reduced from 1.67 to 1.36 (**Supplementary Table 2**), indicating improved stability post-correction.

#### Comet kinetics via Bayesian non-linear regression model

We assume that DNA repair kinetics can be described by two exponential decay processes in parallel: $F(t) = F \cdot e^{-k_f \cdot t} + S \cdot e^{-k_s \cdot t}$, with F≥0, S≥0, $k_f$>0, and $k_s$>0. The sum F+S can be interpreted as the total initial damage; in some data, one phase may dominate if S is negligible or if $k_s$ is extreme. All Bayesian analyses were carried out in R using the "*brms*" package, with our model code and scripts sourced in the Code Availability section.

Parameters F and S (initial damage levels of the fast and slow decay) were assigned weakly informative normal priors centered around tens of percent of DNA in comet tails, based on our prior experience with this assay and replicate controls. The rate constants ($k_f$, $k_s$) were also given weakly informative priors reflecting a plausible range of 0.01-0.05 $min^{-1}$. MCMC was configured with 4 chains, 2000 iterations, 1000 warm-ups, 0.95 adaptative step-size parameter, and a maximum tree depth of 15. We evaluated trace plots for each parameter, and confirmed that the Gelman-Rubin diagnostic (rhat) was below 1.01 for all parameters and checked the effective sample sizes to ensure a thorough exploration of sample space.

Half-lives for the fast phase, slow phase, and overall decay were estimated by solving for the time at which F(t) is half of its initial levels. Specifically, for the overall half-life:

$$F \cdot e^{-k_f t_{\frac{1}{2}}} + S \cdot e^{-k_s t_{\frac{1}{2}}} = \frac{(F+S)}{2} \quad (S0)$$

We used a standard numerical root-finding approach to solve for $t_{1/2}$. Similarly, the fast and slow phase half-lives were computed individually by forcing the exponential term $e^{-k_i t_{1/2}} = 0.5,\ for\ i \in \{f, s\}$.

#### Conceptual framework for variable relationships

Benefited from a comprehensive study design, we considered several important variables contributing to variations in FM-HCR measurements that arise from sample processing and experiment conditions, including batch, CryoTime, CryoAmount, cell viability, and transfection efficiency (TE). These variables were selected based on prior literature [2,3] and our previous applications of FM-HCR in cell line models, which highlighted their potential influence on measurement variability and accuracy. Interactions among these factors, as well as their relationships with individual demographics, were also considered.

To confirm their contributions and explore how these factors interact or mediate the effects of demographics on DRC, we explored the internal relationships among these factors and their external correlations with individual demographics. CryoTime contributed to fluctuations in measurements with mild but significant correlations with several DRC assays (**Supplementary Fig. 3a**), which appeared to reflect its correlation with batch due to the sequential nature of sample collection (**Fig. 1; Supplementary Fig. 3c**) and was thus considered as a batch-related technical factor.

CryoAmount varied by demographics but was relatively consistent across multiple blood draws from the same individual, while TE and viability were influenced by both batch and demographics (**Supplementary Fig. 3c**). After controlling for batch effects and accounting for inter-individual variation, strong associations were observed between TE and MGMT, with an effect estimate expressed in standard deviations of repair capacity [est.] for an interquartile range increase in TE is 0.36 (p = 1.3E-06 [FDR = 1.3E-05]). TE was also associated with LP-BER (est.= 0.17, p = 0.006 [FDR = 0.032]). We furthermore found associations between cell viability and NHEJ (est.=-0.15, p = 0.007 [FDR = 0.023]), HR (est.= −0.14, p = 0.041 [FDR = 0.103]), MMR (est.= 0.17, p = 0.004 [FDR = 0.021]), and NER (est.= 0.18, p = 2.6E-05 [FDR = 2.6E-04]); and between CryoAmount and Hx:T (est.= −0.24, p = 2.8E-04 [FDR = 0.001]), LP-BER (est.= 0.31, p = 1.7E-05 [FDR = 1.7E-04]), A:8oxoG (est.= 0.21, p = 5.3E-05 [FDR = 2.7E-04]). These associations suggest a combination of technical variability and inherent biological influences on DRC (**Fig. 4**), and further explain the remaining within-individual CV in these pathways (**Supplementary Table 1**).

Crude association analysis revealed that individual demographics including age, sex, race, BMI, and smoking status, had influential impacts on DRC pathways (**Fig. 4**). After controlling batch effects and inter-individual variations, significant associations remained. Based on prior knowledge, observed associations, and the temporal relationship among factors, we developed a conceptual framework to guide covariate adjustments in linear mixed-effects models (**Supplementary Fig. 4, DAG1**). The framework highlights the direct and mediated influences of technical and mixed technical-biological factors on DRC, as well as their hypothesized contributions from demographics. These findings underscore the importance of consistent sample handling and the inclusion of relevant sample-specific parameters in downstream analyses to ensure reproducibility and valid inference in population-based DRC studies.

### Linear mixed effects models

Four linear mixed effects (LME) models were applied to assess relationships with DRC.

Model 1 was a null model which included no covariates but individual intercept:

$$Y_{ij} = \beta_0 + \gamma_{0i} + \epsilon_{ij} (S1)$$

This model was compared to a generalized linear model (GLM) without the random intercept component $\gamma_{0i}$ to evaluate inter-individual variations in each DNA repair pathway. A significant likelihood ratio test for the model comparison indicates significantly improved fit by including the random component, thus implying significant inter-individual variations.

Model 2 was a univariate model to evaluate crude associations between each variable and DRC:

$$Y_{ij} = \beta_0 + \boldsymbol{\beta}_1 \boldsymbol{X}_{\boldsymbol{ij}} + \gamma_{0i} + \epsilon_{ij} (S2)$$

Where $\boldsymbol{\beta}_1$ represents the coefficient for variable $\boldsymbol{X}_{\boldsymbol{ij}}$ which might be fixed ($X_{i1} = X_{i2} = \cdots = X_{ij}$) for a given individual i (i.e., the demographics which were either unmodifiable or assumed to remain unchanged within a short follow-up period), or varying for a given individual i across j visits (i.e., the mixed biological-technical factors which were impacted by both demographics and technical conditions in each measurement) (**Supplementary Fig. 4, DAG1**).

Model 3 was a multivariate model to evaluate adjusted associations between demographics and DRC:

$$Y_{ij} = \beta_0 + \boldsymbol{\beta}_1\boldsymbol{X}_i + \gamma_{0i} + \epsilon_{ij}(S3)$$

Where $\boldsymbol{\beta_1}$ is a vector of coefficients for the fixed individual level covariates in vector $\boldsymbol{X}_i$ (age, sex, race, BMI, smoking status) (**Supplementary Fig. 4, DAG2**).

Model 4 was a multivariate model to evaluate adjusted associations between mixed biological-technical factors and DRC:

$$Y_{ij} = \beta_0 + \boldsymbol{\beta}_1\boldsymbol{X}_{ij} + \boldsymbol{\beta}_2\boldsymbol{X}_i + \gamma_{0i} + \epsilon_{ij}(S4)$$

Where $\boldsymbol{\beta_1}$ represents a vector of coefficients for the vector of mixed biological-technical factors $\boldsymbol{X_{ij}}$, and $\boldsymbol{\beta}_2$ represents a vector of coefficients for the vector of fixed individual level demographics $\boldsymbol{X_i}$. We adjusted for the demographics here to control for potential confounding for the relationship between biological-technical factors and DRC (**Supplementary Fig. 4, DAG3**).

### Supplementary Figures

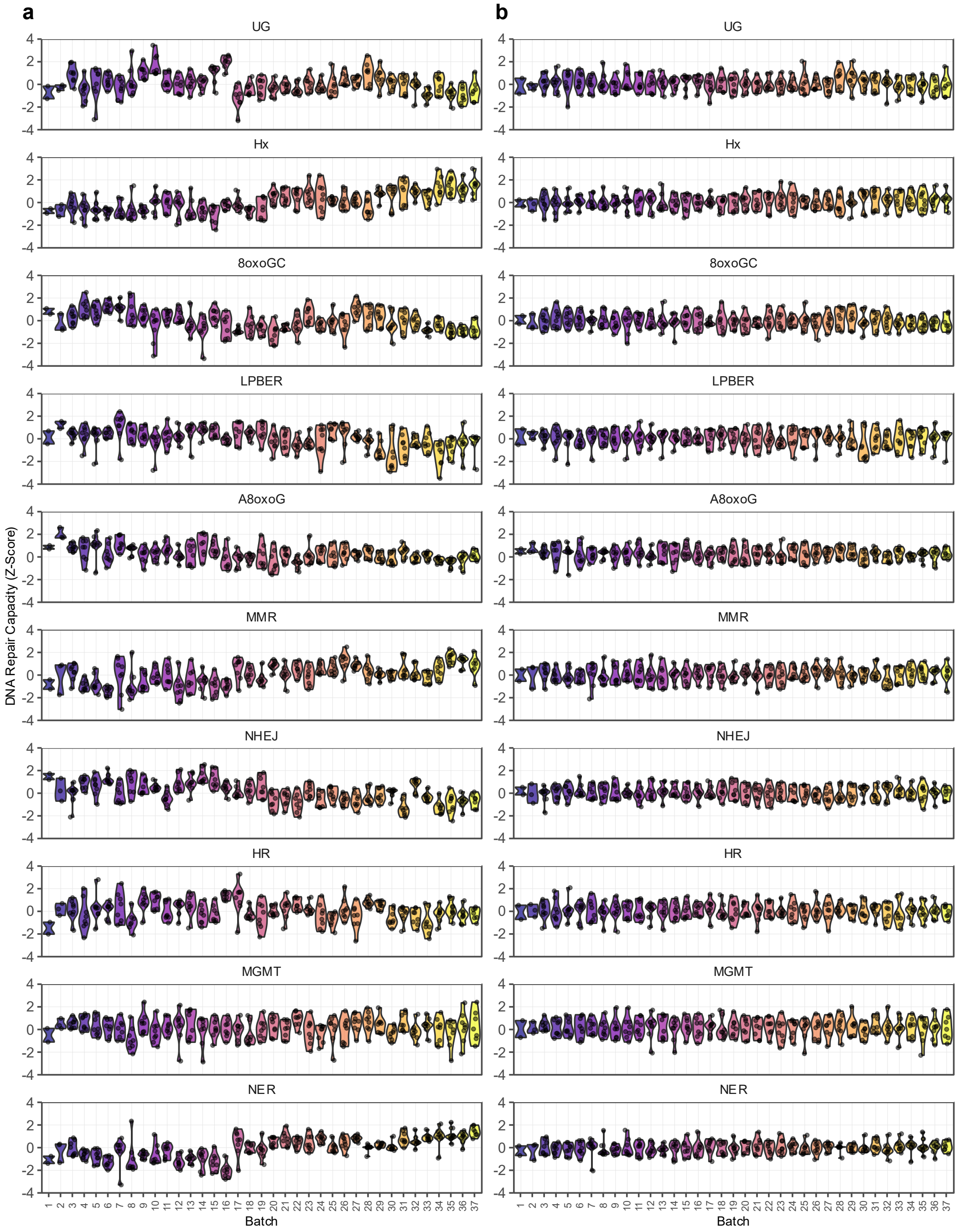


**Supplementary Fig. 1. Standardized DNA repair capacity reported by batch before (a) and after (b) batch correction.** Batch number is reported at the bottom of the plot and a color gradient was applied to facilitate visual comparison of corresponding batches in the two panels and an alternative representation of the same data presented in **Supplementary Fig. 2**. Each data point represents repair capacity for a single technical replicate measurement in one individual's PBMCs. The FM-HCR assay for which data are presented is named at the top of each sub-panel.

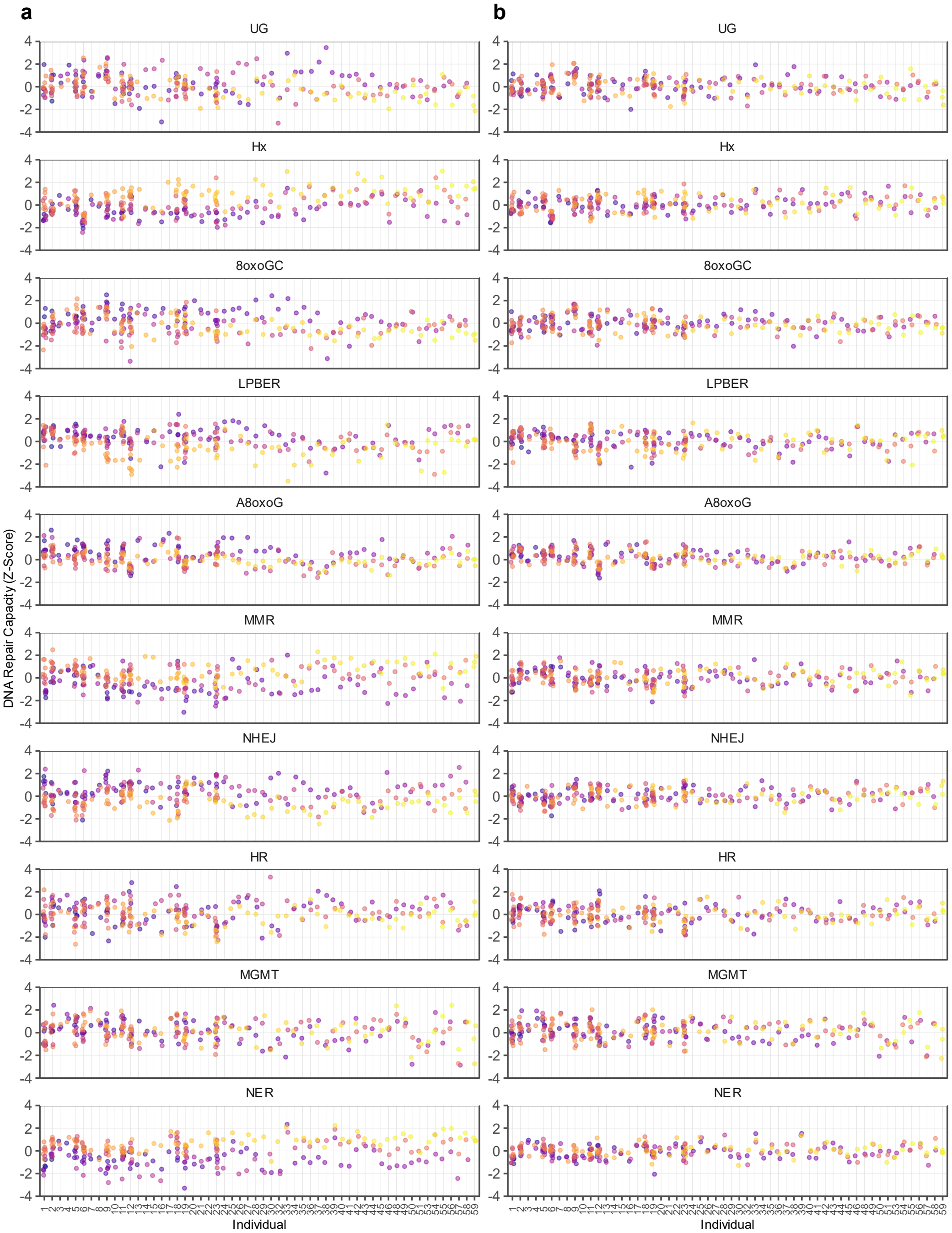


**Supplementary Fig. 2. Standardized DNA repair capacity reported by individual before (a) vs after (b) batch correction.** Individual participant IDs are reported at the bottom of the plot. Each data point represents repair capacity for a single technical replicate measurement in one individual's PBMCs, and the color represents the batch in which the data were collected (See **Supplementary Fig. 1**). Most individuals have 3 technical replicates for PBMCs from a single blood draw; for those with 4-5 repeated measures, up to 15 technical replicates are reported. The FM-HCR assay for which data are presented is named at the top of each sub-panel.

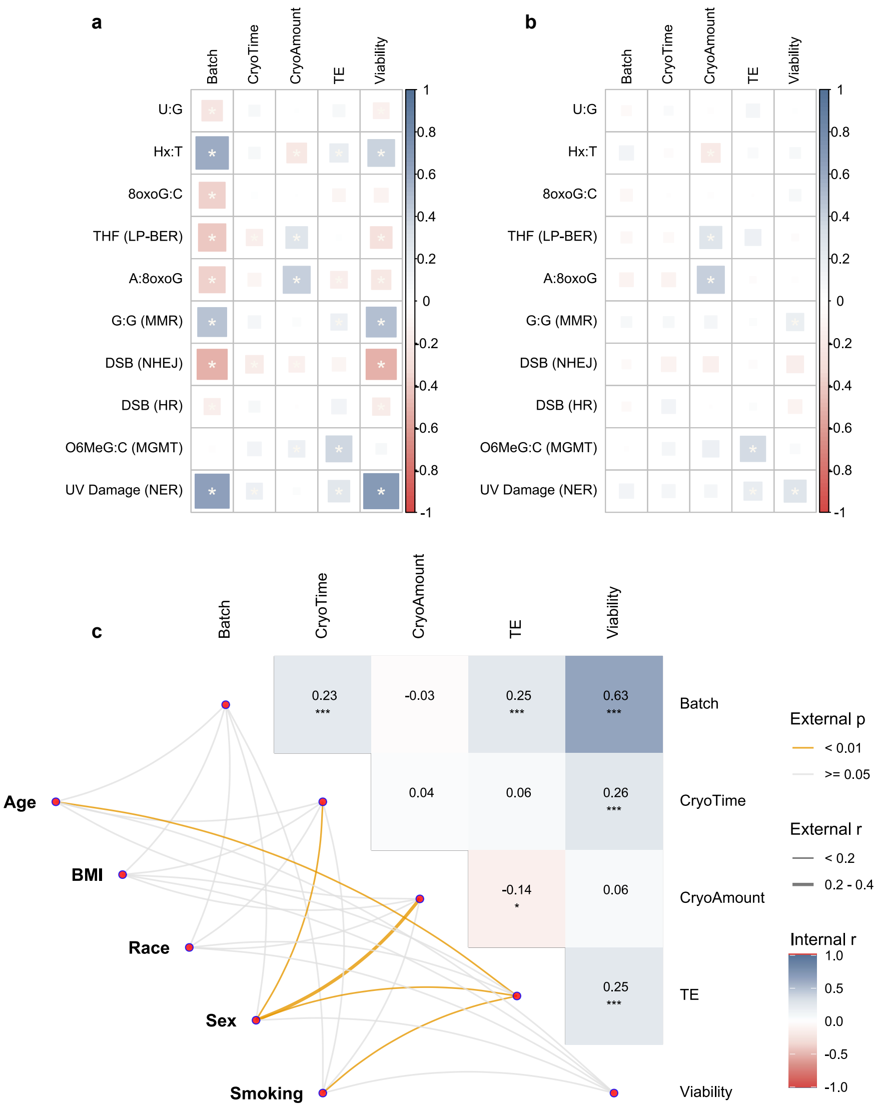


**Supplementary Fig. 3. Relationships among technical factors, biological factors, and DNA repair capacity (DRC). (a)** Spearman's correlations between technical and biological factors and DRC before batch correction. Spearman's r is represented according to the color scale at the right of each plot, and larger boxes represent larger absolute values. (**b**) Spearman's correlations between technical and biological factors and DRC after batch correction. CryoTime, CryoAmount, transfection efficiency (TE), and Viability are defined under study workflow and experimental design in the methods section. **(c)** Internal correlations among technical and biological variables and external correlations between technical and biological variables and demographics. Spearman's r for internal correlations is represented according to the color scale at the right of each plot, and reported explicitly in each box. Spearman's r for external correlations is represented by lines connecting demographic or lifestyle variables at left with the technical and biological variables at right. Thicker lines correspond to stronger correlations and larger absolute values of Spearman's r. Statistically significant correlations ($p < 0.01$) are represented in goldenrod, and non-significant correlations are represented in gray. Multiple testing adjustments were performed using false discovery rate (FDR) at $p < 0.05$.

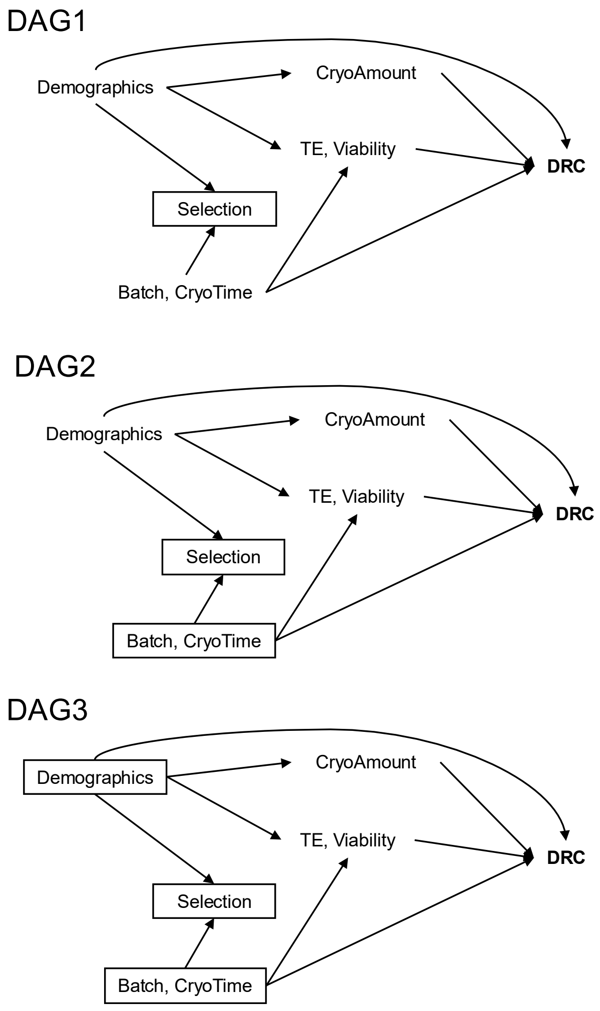


**Supplementary Fig. 4. Directed acyclic graphs (DAGs) illustrating the causal relationships among technical factors, biological factors, and DNA repair capacity (DRC).** **DAG 1**: Temporal and causal relationships among technical factors (batch, CryoTime), mixed biological-technical factors (CryoAmount, cell viability, transfection efficiency [TE]), demographics, and DRC measurements before batch correction. The box around “Selection” describes an observed biased association between demographics and technical factors, which arises due to sample selection. **DAG 2**: This model accounts for technical factors by controlling for batch and CryoTime (indicated by the box around them). It reflects the total effects of demographics on DRC, including both direct effects and indirect effects mediated through CryoAmount, viability, and TE. **DAG 3**: The importance of controlling for demographics when estimating associations between DRC and mixed technical-biological factors (CryoAmount, viability, and TE). Adjusting for demographics and technical factors (as indicated by the boxes) ensures that these estimates are not confounded by the controlled factors.

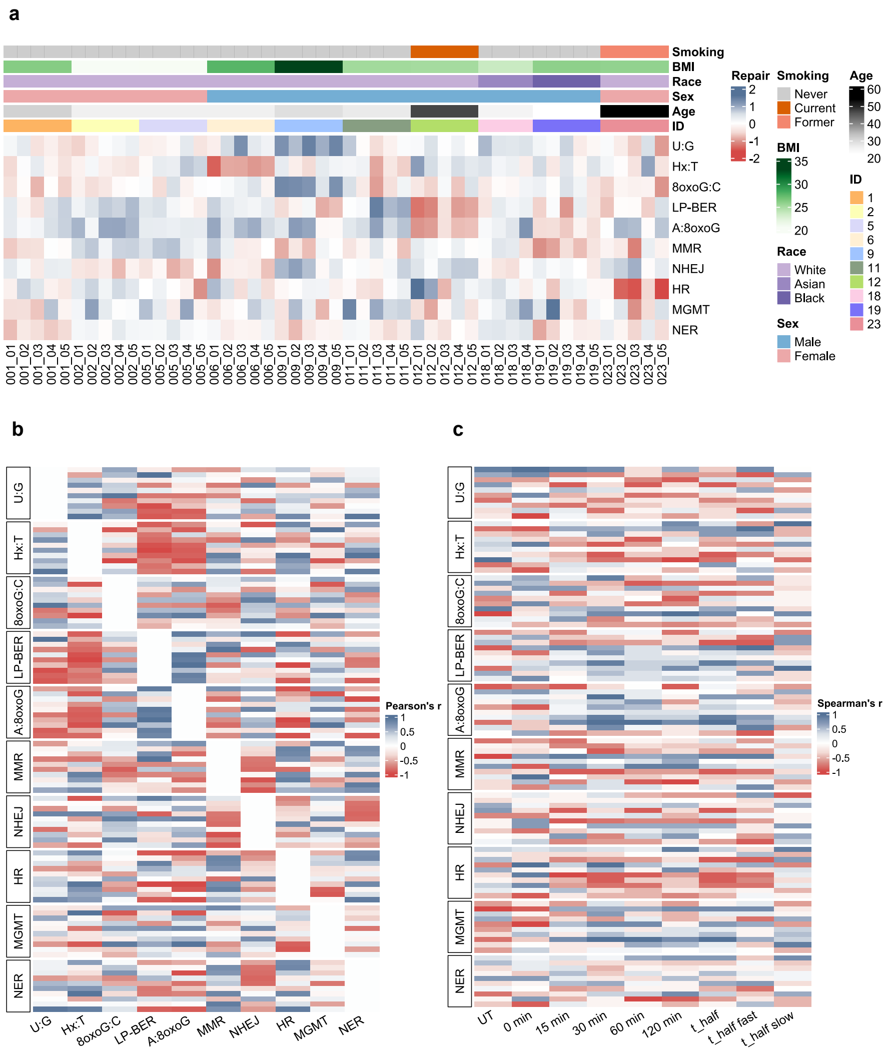


**Supplementary Fig. 5. DNA repair capacity landscape in repeated measures and within-individual correlations. (a)** DNA repair capacity landscape among the 10 individuals with multiple blood draws. Repair capacity was batch-corrected and averaged across replicates for each visit. An alternative representation of the same data presented in Fig. 3b without clustering. **(b)** Heat map representing Pearson's correlation for pairwise correlations between standardized DNA repair capacity (DRC) measured by different FM-HCR assays. Each row represents one of the 10 participants for whom repeated measures were carried out. Colors represent the magnitude and sign of Pearson's r. Each cell in the heatmap represents the correlation between 4 or 5 biological replicates, each of which was calculated from the average of three technical replicates. **(c)** Heat map representing Spearman's r for pairwise correlations between CometChip parameters and DRC measured by different FM-HCR assays. Each row represents one of the 10 participants for whom repeated measures were carried out. Colors represent the magnitude and sign of Spearman's r. Each cell in the heatmap represents the correlation between 4 or 5 biological replicates, each of which was calculated from the average of three technical replicates.

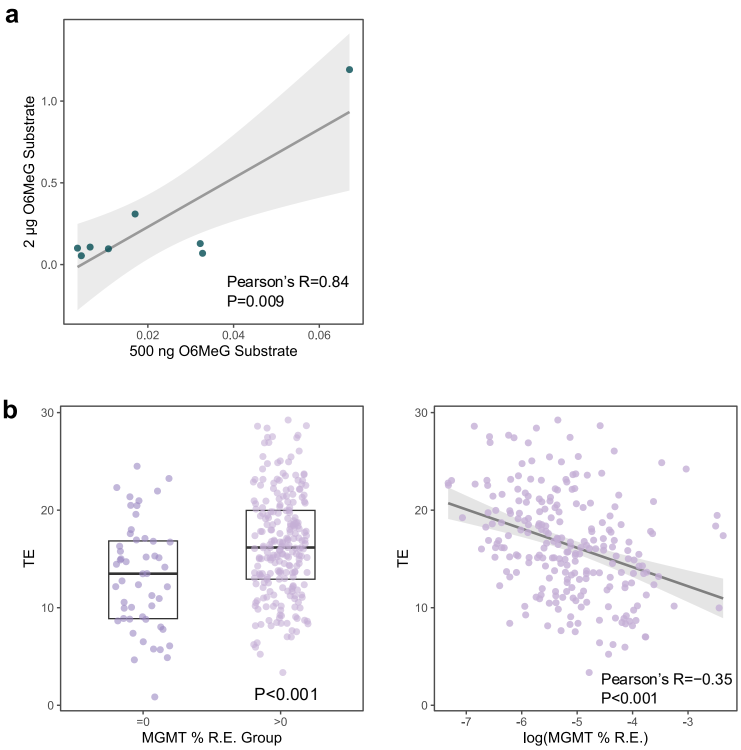


**Supplementary Fig. 6. Associations between transfection efficiency (TE) and MGMT reporter expression. (a)** Strong positive correlations between reporter expression in stimulated lymphocytes with 500 ng vs 2 μg of reporter plasmid. **(b)** Stimulated lymphocytes exhibited an inverse relationship between TE and MGMT reporter expression. The box and whisker plot (left) illustrates that TE was lower among samples for which the MGMT assay yielded zero fluorescence positive cells. The correlation plot at right illustrates that samples with higher TE exhibited lower fluorescent reporter expression, corresponding to higher MGMT activity.

### Supplementary Tables

Supplementary Table 1. Batch effect correction evaluation.

Supplementary Table 2. Within-individual stability metrics before and after batch correction.

Supplementary Table 3. Associations between batch-corrected DNA repair capacity with individual demographics and biological factors after multiple imputation.

Supplementary Table 4. Across-individual 10-pathway distances.

Supplementary Table 5. Improved model fitting accuracy in repeated measures (CV%).

Supplementary Table 6. Individual differences in Comet kinetics.

Supplementary Table 7. Association between transfection efficiency (TE) and MGMT repair capacity.

Supplementary Table 8. Interindividual variations in DNA repair capacity from previous literature.

Supplementary Table 9. Plasmids for FM-HCR.

**Supplementary Table 1.** Batch effect correction evaluation

Supplementary Table 1.a. Batch effect correction using experimental-sample only methods.

| Coefficient of variation | UG | Hx | 8oxoGC | LP-BER | A8oxoG | MMR | NHEJ | HR | MGMT | NER | Summary | % reduction |
|---|---|---|---|---|---|---|---|---|---|---|---|---|
| *Across-batch CV (among batch-averages across 37 batches)* | | | | | | | | | | | | |
| Before batch correction | 26.4 | 18.7 | 13.5 | 12.4 | 34.5 | 19.7 | 14.8 | 40.6 | 45.7 | 31.8 | 25.8 | |
| Post batch correction | 6.7 | 3.3 | 2.9 | 3.3 | 14.4 | 4.3 | 2.7 | 6.5 | 18.2 | 5.0 | 6.7 | -74.0 |
| *Within-individual CV (among replicates of same individual)* | | | | | | | | | | | | |
| Before batch correction | 31.2 | 20.3 | 16.2 | 15.9 | 36.5 | 24.0 | 16.1 | 45.7 | 46.5 | 34.0 | 28.6 | |
| Post batch correction | 20.9 | 12.1 | 10.1 | 10.8 | 24.5 | 14.3 | 9.0 | 32.6 | 46.7 | 15.1 | 19.6 | -31.5 |

Supplementary Table 1.b. Batch effect corretion using experimental samples plus the control.

| Coefficient of variation | UG | Hx | 8oxoGC | LP-BER | A8oxoG | MMR | NHEJ | HR | MGMT | NER | Summary | % reduction |
|---|---|---|---|---|---|---|---|---|---|---|---|---|
| *Across-batch CV* | | | | | | | | | | | | |
| Before batch correction | 26.4 | 18.7 | 13.5 | 12.4 | 34.5 | 19.7 | 14.8 | 40.6 | 45.7 | 31.8 | 25.8 | |
| Post batch correction | 7.6 | 5.0 | 3.4 | 4.5 | 17.4 | 4.7 | 3.6 | 11.1 | 27.9 | 7.5 | 9.3 | -64.1 |
| *Within-individual CV* | | | | | | | | | | | | |
| Before batch correction | 31.2 | 20.3 | 16.2 | 15.9 | 36.5 | 24.0 | 16.1 | 45.7 | 46.5 | 34.0 | 28.6 | |
| Post batch correction | 21.1 | 12.2 | 10.4 | 11.1 | 26.5 | 14.7 | 9.4 | 34.2 | 47.1 | 15.9 | 20.2 | -29.3 |

Supplementary Table 1.c. Batch effect corretion in the control samples.

| Coefficient of variation | UG | Hx | 8oxoGC | LP-BER | A8oxoG | MMR | NHEJ | HR | MGMT | NER | Summary | % reduction |
|---|---|---|---|---|---|---|---|---|---|---|---|---|
| *#000 corrected with experimental samples* | | | | | | | | | | | | |
| Before batch correction | 30.2 | 17.8 | 24.7 | 19.5 | 34.6 | 21.8 | 25.7 | 57.4 | 98.3 | 28.0 | 35.8 | |
| Post batch correction | 19.2 | 11.3 | 13.7 | 14.8 | 31.3 | 18.3 | 13.8 | 39.2 | 56.5 | 23.7 | 24.2 | -32.5 |

**Supplementary Table 2.** Within-individual stability metrics before and after batch correction

| Averaged Coefficient of Variation | | | Averaged 10-pathway distances | | |
|---|---|---|---|---|---|
| **Pathway** | **Before** | **After** | **ID** | **Before** | **After** |
| UG | 18.2 | 14.8 | 1 | 1.70 | 1.30 |
| Hx | 9.1 | 9.9 | 2 | 1.33 | 0.93 |
| 8oxoGC | 10.4 | 7.4 | 5 | 1.40 | 1.23 |
| LP-BER | 8.4 | 8.2 | 6 | 1.57 | 1.30 |
| A8oxoG | 27.2 | 23.8 | 9 | 1.91 | 1.46 |
| MMR | 13.8 | 10.1 | 11 | 1.69 | 1.54 |
| NHEJ | 8.2 | 6.0 | 12 | 1.62 | 1.51 |
| HR | 37.4 | 28.9 | 18 | 1.67 | 1.04 |
| MGMT | 53.1 | 47.9 | 19 | 1.85 | 1.49 |
| NER | 14.8 | 10.2 | 23 | 1.97 | 1.82 |
| Average | 20.1 | 16.7 | Average | 1.67 | 1.36 |

**Supplementary Table 3**. Associations between batch-corrected DNA repair capacity with individual demographics and biological factors after multiple imputation.

| **Crude** | **U:G** | | | **Hx:T** | | | **8oxoG:C** | | | **LP-BER** | | | **A:8oxoG** | | | **MMR** | | | **NHEJ** | | | **HR** | | | **MGMT** | | | **NER** | | |
|---|---|---|---|---|---|---|---|---|---|---|---|---|---|---|---|---|---|---|---|---|---|---|---|---|---|---|---|---|---|---|
| | **beta** | **p-value** | **fdr** | **beta** | **p-value** | **fdr** | **beta** | **p-value** | **fdr** | **beta** | **p-value** | **fdr** | **beta** | **p-value** | **fdr** | **beta** | **p-value** | **fdr** | **beta** | **p-value** | **fdr** | **beta** | **p-value** | **fdr** | **beta** | **p-value** | **fdr** | **beta** | **p-value** | **fdr** |
| **Age (years)** | -0.012 | *0.032* | ***0.098*** | 0.006 | 0.567 | 0.631 | -0.012 | *0.040* | ***0.098*** | -0.008 | 0.302 | 0.504 | -0.010 | *0.049* | ***0.098*** | 0.006 | 0.435 | 0.622 | 0.010 | *0.030* | ***0.098*** | -0.001 | 0.720 | 0.720 | -0.014 | *0.021* | ***0.098*** | 0.005 | 0.509 | 0.631 |
| **Sex (female vs male)** | **-0.398** | *0.001* | ***0.010*** | 0.101 | 0.469 | 0.700 | -0.265 | *0.023* | 0.117 | 0.102 | 0.490 | 0.700 | 0.044 | 0.719 | 0.760 | 0.218 | 0.097 | 0.323 | -0.035 | 0.760 | 0.760 | 0.186 | 0.136 | 0.339 | -0.128 | 0.462 | 0.700 | -0.057 | 0.649 | 0.760 |
| **BMI (kg/m2)** | 0.035 | *0.032* | 0.255 | 0.010 | 0.441 | 0.630 | 0.024 | 0.258 | 0.551 | -0.035 | 0.076 | 0.255 | 0.011 | 0.668 | 0.743 | -0.028 | 0.077 | 0.255 | 0.011 | 0.339 | 0.565 | -0.005 | 0.597 | 0.743 | 0.002 | 0.776 | 0.776 | 0.013 | 0.275 | 0.551 |
| **Race** | | | | | | | | | | | | | | | | | | | | | | | | | | | | | | |
| White (reference) | - | - | | - | - | | - | - | | - | - | | - | - | | - | - | | - | - | | - | - | | - | - | | - | - | |
| Asian | 0.091 | 0.590 | 0.684 | 0.370 | *0.026* | ***0.088*** | -0.094 | 0.544 | 0.684 | -0.093 | 0.616 | 0.684 | -0.055 | 0.708 | 0.708 | 0.293 | *0.050* | 0.125 | -0.072 | 0.613 | 0.684 | 0.186 | 0.236 | 0.471 | **0.585** | *0.004* | ***0.040*** | 0.337 | *0.021* | ***0.088*** |
| Black/African American | -0.025 | 0.931 | 0.944 | -0.262 | 0.369 | 0.737 | 0.019 | 0.944 | 0.944 | 0.171 | 0.597 | 0.853 | -0.552 | *0.033* | 0.165 | **-0.782** | *0.002* | ***0.022*** | 0.399 | 0.107 | 0.355 | -0.139 | 0.597 | 0.853 | 0.139 | 0.694 | 0.867 | -0.345 | 0.180 | 0.451 |
| **Smoking status** | | | | | | | | | | | | | | | | | | | | | | | | | | | | | | |
| Never (reference) | - | - | | - | - | | - | - | | - | - | | - | - | | - | - | | - | - | | - | - | | - | - | | - | - | |
| Current | -0.231 | 0.491 | 0.737 | -0.113 | 0.752 | 0.835 | 0.163 | 0.589 | 0.737 | -0.798 | *0.023* | 0.230 | -0.199 | 0.536 | 0.737 | -0.260 | 0.456 | 0.737 | 0.443 | 0.103 | 0.434 | -0.017 | 0.956 | 0.956 | -0.660 | 0.130 | 0.434 | -0.278 | 0.393 | 0.737 |
| Former | -0.273 | 0.157 | 0.365 | 0.293 | 0.143 | 0.365 | -0.278 | 0.114 | 0.365 | -0.174 | 0.391 | 0.558 | -0.029 | 0.869 | 0.914 | -0.021 | 0.914 | 0.914 | 0.344 | *0.030* | 0.301 | -0.054 | 0.767 | 0.914 | -0.240 | 0.329 | 0.549 | 0.238 | 0.188 | 0.365 |
| **CryoAmount (millons per** | -0.013 | 0.282 | 0.470 | **-0.034** | *0.004* | ***0.012*** | 0.002 | 0.826 | 0.836 | **0.045** | *0.000* | ***0.001*** | **0.041** | *8.0E-06* | **######** | 0.004 | 0.732 | 0.836 | -0.006 | 0.584 | 0.834 | -0.002 | 0.836 | 0.836 | 0.026 | 0.092 | 0.230 | -0.011 | 0.281 | 0.470 |
| **Transfection efficiency** | 0.014 | 0.082 | 0.163 | 0.009 | 0.215 | 0.358 | -0.003 | 0.738 | 0.763 | 0.018 | *0.030* | ***0.076*** | -0.013 | *0.030* | ***0.076*** | 0.002 | 0.763 | 0.763 | 0.006 | 0.367 | 0.524 | -0.004 | 0.676 | 0.763 | **0.043** | *1.4E-06* | **1.4E-05** | **0.019** | *0.001* | ***0.007*** |
| **Cell viability** | -0.002 | 0.711 | 0.789 | 0.002 | 0.565 | 0.789 | 0.005 | 0.225 | 0.449 | -0.002 | 0.664 | 0.789 | -0.002 | 0.480 | 0.789 | **0.012** | *0.003* | ***0.014*** | -0.009 | *0.020* | ***0.066*** | -0.010 | *0.041* | 0.101 | 0.000 | 0.949 | 0.949 | **0.014** | *2.2E-06* | ***2.2E-05*** |

| **Multivariate - demograph** | **UG** | | | **Hx** | | | **8oxoGC** | | | **LP-BER** | | | **A8oxoG** | | | **MMR** | | | **NHEJ** | | | **HR** | | | **MGMT** | | | **NER** | | |
|---|---|---|---|---|---|---|---|---|---|---|---|---|---|---|---|---|---|---|---|---|---|---|---|---|---|---|---|---|---|---|
| | **beta** | **p-value** | **fdr** | **beta** | **p-value** | **fdr** | **beta** | **p-value** | **fdr** | **beta** | **p-value** | **fdr** | **beta** | **p-value** | **fdr** | **beta** | **p-value** | **fdr** | **beta** | **p-value** | **fdr** | **beta** | **p-value** | **fdr** | **beta** | **p-value** | **fdr** | **beta** | **p-value** | **fdr** |
| **Age (years)** | -0.011 | 0.197 | 0.395 | 0.003 | 0.980 | 0.980 | -0.015 | 0.132 | 0.395 | -0.004 | 0.629 | 0.786 | **-0.021** | *0.004* | ***0.037*** | 0.016 | 0.171 | 0.395 | 0.005 | 0.302 | 0.503 | 0.000 | 0.818 | 0.908 | -0.008 | 0.178 | 0.395 | 0.006 | 0.626 | 0.786 |
| **Sex (female vs male)** | -0.302 | 0.060 | 0.457 | 0.115 | 0.408 | 0.794 | -0.130 | 0.315 | 0.794 | -0.067 | 0.556 | 0.794 | 0.110 | 0.444 | 0.794 | 0.073 | 0.688 | 0.860 | -0.019 | 0.903 | 0.903 | 0.262 | 0.091 | 0.457 | -0.079 | 0.820 | 0.903 | -0.110 | 0.493 | 0.794 |
| **BMI (kg/m2)** | 0.033 | 0.052 | 0.376 | 0.015 | 0.445 | 0.741 | 0.028 | 0.237 | 0.593 | -0.039 | 0.077 | 0.376 | 0.024 | 0.394 | 0.741 | -0.027 | 0.113 | 0.376 | 0.001 | 0.798 | 0.798 | 0.013 | 0.650 | 0.751 | 0.012 | 0.598 | 0.751 | 0.005 | 0.676 | 0.751 |
| **Race** | | | | | | | | | | | | | | | | | | | | | | | | | | | | | | |
| White (reference) | - | - | | - | - | | - | - | | - | - | | - | - | | - | - | | - | - | | - | - | | - | - | | - | - | |
| Asian | 0.003 | 0.918 | 0.918 | **0.441** | *0.020* | ***0.098*** | -0.172 | 0.220 | 0.325 | -0.232 | 0.263 | 0.328 | -0.153 | 0.209 | 0.325 | 0.337 | *0.029* | ***0.098*** | 0.010 | 0.840 | 0.918 | 0.233 | 0.228 | 0.325 | 0.499 | *0.043* | 0.108 | 0.376 | *0.021* | ***0.098*** |
| Black/African American | -0.233 | 0.365 | 0.730 | -0.169 | 0.553 | 0.854 | -0.135 | 0.598 | 0.854 | 0.047 | 0.882 | 0.961 | **-0.651** | *0.007* | ***0.037*** | **-0.699** | *0.005* | ***0.037*** | 0.496 | *0.036* | 0.119 | -0.067 | 0.768 | 0.960 | 0.017 | 0.961 | 0.961 | -0.317 | 0.223 | 0.558 |
| **Smoking status** | | | | | | | | | | | | | | | | | | | | | | | | | | | | | | |
| Never (reference) | - | - | | - | - | | - | - | | - | - | | - | - | | - | - | | - | - | | - | - | | - | - | | - | - | |
| Current | -0.266 | 0.405 | 0.675 | 0.015 | 0.905 | 0.905 | 0.195 | 0.631 | 0.789 | -0.834 | *0.026* | 0.258 | -0.037 | 0.829 | 0.905 | -0.338 | 0.356 | 0.675 | 0.422 | 0.186 | 0.675 | 0.154 | 0.614 | 0.789 | -0.499 | 0.357 | 0.675 | -0.331 | 0.365 | 0.675 |
| Former | -0.070 | 0.515 | 0.772 | 0.203 | 0.395 | 0.772 | -0.017 | 0.624 | 0.772 | 0.019 | 0.754 | 0.772 | 0.279 | 0.244 | 0.772 | -0.345 | 0.449 | 0.772 | 0.282 | 0.352 | 0.772 | -0.138 | 0.729 | 0.772 | -0.014 | 0.772 | 0.772 | 0.133 | 0.472 | 0.772 |

| **Multivariate - mixed** | **UG** | | | **Hx** | | | **8oxoGC** | | | **LP-BER** | | | **A8oxoG** | | | **MMR** | | | **NHEJ** | | | **HR** | | | **MGMT** | | | **NER** | | |
|---|---|---|---|---|---|---|---|---|---|---|---|---|---|---|---|---|---|---|---|---|---|---|---|---|---|---|---|---|---|---|
| | **beta** | **p-value** | **fdr** | **beta** | **p-value** | **fdr** | **beta** | **p-value** | **fdr** | **beta** | **p-value** | **fdr** | **beta** | **p-value** | **fdr** | **beta** | **p-value** | **fdr** | **beta** | **p-value** | **fdr** | **beta** | **p-value** | **fdr** | **beta** | **p-value** | **fdr** | **beta** | **p-value** | **fdr** |
| **CryoAmount (millons per** | -0.009 | 0.485 | 0.693 | **-0.043** | *2.8E-04* | ***0.001*** | 0.005 | 0.593 | 0.742 | **0.055** | *1.7E-05* | **1.7E-04** | **0.037** | *5.3E-05* | **######** | -0.002 | 0.795 | 0.883 | 0.000 | 0.954 | 0.954 | -0.010 | 0.415 | 0.692 | 0.030 | 0.072 | 0.181 | -0.012 | 0.212 | 0.424 |
| **Transfection efficiency** | 0.010 | 0.246 | 0.411 | 0.006 | 0.459 | 0.574 | -0.008 | 0.329 | 0.470 | **0.023** | *0.006* | ***0.032*** | -0.010 | 0.081 | 0.161 | -0.003 | 0.651 | 0.723 | 0.014 | *0.045* | 0.149 | 0.002 | 0.911 | 0.911 | **0.047** | *1.3E-06* | **1.3E-05** | 0.011 | 0.079 | 0.161 |
| **Cell viability** | -0.002 | 0.666 | 0.740 | 0.002 | 0.648 | 0.740 | 0.007 | 0.103 | 0.206 | -0.007 | 0.126 | 0.210 | 0.000 | 0.911 | 0.911 | **0.012** | *0.004* | ***0.021*** | **-0.011** | *0.007* | ***0.023*** | -0.010 | *0.041* | 0.103 | -0.007 | 0.169 | 0.241 | **0.013** | *2.6E-05* | ***2.6E-04*** |

Three linear mixed-effect (LME) models were fit to explore relationships with measured DNA repair capacity: crude model; multivaraite model on individual level demographics; multivariate model for mixed technical-biological factors.

Note: beta values presented here correspond to effect size of each variable by 1 unit increase.

The effect esitmates were obtained by pooling multiple imputation sets for missing values.

Both the p values (p-value) from the original LME models and the multiple-testing corrected p values (fdr) are presented.

BMI: body mass index; CryoAmount: cell amount available for cryopreservation after isolation; fdr: false discovery rate.

**Associations between batch-corrected DNA repair capacity with individual demographics and biological factors using complete case analysis.**

| **Crude** | **U:G** | | | **Hx:T** | | | **8oxoG:C** | | | **LP-BER** | | | **A:8oxoG** | | | **MMR** | | | **NHEJ** | | | **HR** | | | **MGMT** | | | **NER** | | |
|---|---|---|---|---|---|---|---|---|---|---|---|---|---|---|---|---|---|---|---|---|---|---|---|---|---|---|---|---|---|---|
| | **beta** | **p-value** | **fdr** | **beta** | **p-value** | **fdr** | **beta** | **p-value** | **fdr** | **beta** | **p-value** | **fdr** | **beta** | **p-value** | **fdr** | **beta** | **p-value** | **fdr** | **beta** | **p-value** | **fdr** | **beta** | **p-value** | **fdr** | **beta** | **p-value** | **fdr** | **beta** | **p-value** | **fdr** |
| **Age (years)** | -0.011 | 0.057 | 0.144 | 0.006 | 0.324 | 0.414 | **-0.011** | *0.040* | 0.144 | -0.008 | 0.200 | 0.378 | -0.010 | 0.063 | 0.144 | 0.007 | 0.266 | 0.409 | 0.010 | 0.063 | 0.144 | 0.000 | 0.947 | 0.915 | -0.014 | 0.070 | 0.144 | 0.006 | 0.272 | 0.409 |
| **Sex (female vs male)** | **-0.398** | *0.002* | ***0.020*** | 0.101 | 0.473 | 0.704 | **-0.265** | *0.028* | 0.140 | 0.102 | 0.493 | 0.704 | 0.044 | 0.720 | 0.761 | 0.218 | 0.103 | 0.343 | -0.035 | 0.761 | 0.761 | 0.186 | 0.141 | 0.353 | -0.128 | 0.466 | 0.704 | -0.057 | 0.651 | 0.761 |
| **BMI (kg/m2)** | **0.042** | *0.020* | 0.315 | 0.010 | 0.619 | 0.736 | 0.028 | 0.093 | 0.330 | -0.039 | 0.054 | 0.315 | 0.014 | 0.408 | 0.707 | -0.032 | 0.081 | 0.317 | 0.007 | 0.666 | 0.707 | -0.008 | 0.648 | 0.833 | 0.006 | 0.794 | 0.951 | 0.014 | 0.438 | 0.707 |
| **Race** | | | | | | | | | | | | | | | | | | | | | | | | | | | | | | |
| White (reference) | - | - | | - | - | | - | - | | - | - | | - | - | | - | - | | - | - | | - | - | | - | - | | - | - | |
| Asian | 0.091 | 0.592 | 0.686 | **0.370** | *0.030* | 0.100 | -0.094 | 0.546 | 0.686 | -0.093 | 0.617 | 0.686 | -0.055 | 0.710 | 0.710 | 0.293 | 0.055 | 0.138 | -0.072 | 0.615 | 0.686 | 0.186 | 0.240 | 0.480 | **0.585** | *0.006* | ***0.060*** | **0.337** | *0.025* | 0.100 |
| Black/African Americar | -0.025 | 0.932 | 0.945 | -0.262 | 0.373 | 0.746 | 0.019 | 0.945 | 0.945 | 0.171 | 0.600 | 0.857 | **-0.552** | *0.038* | 0.190 | **-0.782** | *0.004* | ***0.040*** | 0.399 | 0.113 | 0.377 | -0.139 | 0.600 | 0.857 | 0.139 | 0.696 | 0.870 | -0.345 | 0.187 | 0.468 |
| **Smoking status** | | | | | | | | | | | | | | | | | | | | | | | | | | | | | | |
| Never (reference) | - | - | | - | - | | - | - | | - | - | | - | - | | - | - | | - | - | | - | - | | - | - | | - | - | |
| Current | -0.231 | 0.495 | 0.741 | -0.113 | 0.753 | 0.837 | 0.163 | 0.593 | 0.741 | **-0.798** | *0.029* | 0.290 | -0.199 | 0.536 | 0.741 | -0.260 | 0.456 | 0.741 | 0.443 | 0.112 | 0.463 | -0.017 | 0.956 | 0.956 | -0.660 | 0.139 | 0.463 | -0.278 | 0.393 | 0.741 |
| Former | -0.273 | 0.162 | 0.376 | 0.293 | 0.148 | 0.376 | -0.278 | 0.119 | 0.376 | -0.174 | 0.395 | 0.564 | -0.029 | 0.869 | 0.914 | -0.021 | 0.914 | 0.914 | **0.344** | *0.034* | 0.340 | -0.054 | 0.768 | 0.914 | -0.240 | 0.334 | 0.557 | 0.238 | 0.188 | 0.376 |
| **CryoAmount (millons per** | -0.013 | 0.262 | 0.472 | **-0.035** | *0.003* | ***0.013*** | 0.003 | 0.819 | 0.831 | **0.046** | **<0.001** | **<0.001** | **0.041** | **<0.001** | ***0.000*** | 0.003 | 0.779 | 0.831 | -0.005 | 0.647 | 0.831 | -0.003 | 0.772 | 0.831 | 0.023 | 0.112 | 0.190 | -0.012 | 0.249 | 0.472 |
| **Transfection efficiency** | 0.014 | 0.083 | 0.166 | 0.009 | 0.216 | 0.360 | -0.003 | 0.739 | 0.764 | **0.018** | *0.031* | ***0.078*** | **-0.013** | *0.031* | ***0.078*** | 0.002 | 0.764 | 0.764 | 0.006 | 0.367 | 0.524 | -0.004 | 0.676 | 0.764 | **0.043** | **<0.001** | ***2.3E-05*** | **0.019** | *0.002* | ***0.010*** |
| **Cell viability** | -0.002 | 0.711 | 0.790 | 0.002 | 0.566 | 0.790 | 0.005 | 0.226 | 0.452 | -0.002 | 0.664 | 0.790 | -0.002 | 0.481 | 0.790 | **0.012** | *0.003* | ***0.015*** | **-0.009** | *0.020* | ***0.067*** | **-0.010** | *0.042* | 0.105 | 0.000 | 0.949 | 0.949 | **0.014** | **<0.001** | ***3.7E-05*** |

| **Multivariate - demograph** | **UG** | | | **Hx** | | | **8oxoGC** | | | **LP-BER** | | | **A8oxoG** | | | **MMR** | | | **NHEJ** | | | **HR** | | | **MGMT** | | | **NER** | | |
|---|---|---|---|---|---|---|---|---|---|---|---|---|---|---|---|---|---|---|---|---|---|---|---|---|---|---|---|---|---|---|
| | **beta** | **p-value** | **fdr** | **beta** | **p-value** | **fdr** | **beta** | **p-value** | **fdr** | **beta** | **p-value** | **fdr** | **beta** | **p-value** | **fdr** | **beta** | **p-value** | **fdr** | **beta** | **p-value** | **fdr** | **beta** | **p-value** | **fdr** | **beta** | **p-value** | **fdr** | **beta** | **p-value** | **fdr** |
| **Age (years)** | -0.010 | 0.195 | 0.370 | 0.003 | 0.692 | 0.847 | -0.014 | 0.067 | 0.140 | -0.005 | 0.569 | 0.825 | **-0.022** | *0.003* | ***0.030*** | **0.017** | *0.027* | 0.140 | 0.004 | 0.547 | 0.733 | 0.001 | 0.878 | 0.959 | -0.009 | 0.400 | 0.675 | 0.007 | 0.352 | 0.675 |
| **Sex (female vs male)** | **-0.297** | *0.039* | 0.430 | 0.122 | 0.448 | 0.775 | -0.135 | 0.326 | 0.775 | -0.081 | 0.625 | 0.782 | 0.112 | 0.392 | 0.775 | 0.072 | 0.599 | 0.782 | -0.026 | 0.843 | 0.887 | 0.251 | 0.105 | 0.440 | -0.066 | 0.740 | 0.782 | -0.105 | 0.455 | 0.775 |
| **BMI (kg/m2)** | 0.036 | 0.061 | 0.286 | 0.017 | 0.428 | 0.694 | 0.028 | 0.122 | 0.286 | **-0.044** | *0.050* | 0.286 | 0.027 | 0.132 | 0.286 | -0.028 | 0.128 | 0.286 | -0.002 | 0.927 | 0.946 | 0.010 | 0.608 | 0.694 | 0.017 | 0.514 | 0.780 | 0.007 | 0.701 | 0.851 |
| **Race** | | | | | | | | | | | | | | | | | | | | | | | | | | | | | | |
| White (reference) | - | - | | - | - | | - | - | | - | - | | - | - | | - | - | | - | - | | - | - | | - | - | | - | - | |
| Asian | 0.021 | 0.893 | 0.976 | **0.448** | *0.015* | ***0.080*** | -0.157 | 0.305 | 0.364 | -0.246 | 0.185 | 0.345 | -0.141 | 0.335 | 0.364 | **0.327** | *0.036* | ***0.080*** | 0.001 | 0.992 | 0.976 | 0.232 | 0.177 | 0.345 | **0.512** | *0.024* | ***0.080*** | **0.380** | *0.018* | ***0.080*** |
| Black/African Americar | -0.226 | 0.385 | 0.740 | -0.163 | 0.593 | 0.840 | -0.129 | 0.609 | 0.840 | 0.036 | 0.906 | 0.965 | **-0.652** | *0.011* | ***0.060*** | **-0.697** | *0.009* | ***0.060*** | 0.494 | 0.050 | 0.160 | -0.065 | 0.816 | 0.965 | 0.020 | 0.957 | 0.965 | -0.311 | 0.248 | 0.608 |
| **Smoking status** | | | | | | | | | | | | | | | | | | | | | | | | | | | | | | |
| Never (reference) | - | - | | - | - | | - | - | | - | - | | - | - | | - | - | | - | - | | - | - | | - | - | | - | - | |
| Current | -0.265 | 0.404 | 0.713 | 0.017 | 0.964 | 0.958 | 0.182 | 0.555 | 0.727 | **-0.840** | *0.032* | 0.330 | -0.031 | 0.920 | 0.958 | -0.345 | 0.269 | 0.646 | 0.417 | 0.172 | 0.646 | 0.139 | 0.685 | 0.809 | -0.482 | 0.298 | 0.646 | -0.331 | 0.321 | 0.646 |
| Former | -0.070 | 0.766 | 0.985 | 0.199 | 0.456 | 0.972 | -0.029 | 0.898 | 0.985 | 0.012 | 0.965 | 0.985 | 0.299 | 0.168 | 0.693 | -0.372 | 0.107 | 0.693 | 0.295 | 0.182 | 0.693 | -0.138 | 0.589 | 0.972 | -0.008 | 0.981 | 0.985 | 0.119 | 0.608 | 0.972 |

| **Multivariate - mixed** | **UG** | | | **Hx** | | | **8oxoGC** | | | **LP-BER** | | | **A8oxoG** | | | **MMR** | | | **NHEJ** | | | **HR** | | | **MGMT** | | | **NER** | | |
|---|---|---|---|---|---|---|---|---|---|---|---|---|---|---|---|---|---|---|---|---|---|---|---|---|---|---|---|---|---|---|
| | **beta** | **p-value** | **fdr** | **beta** | **p-value** | **fdr** | **beta** | **p-value** | **fdr** | **beta** | **p-value** | **fdr** | **beta** | **p-value** | **fdr** | **beta** | **p-value** | **fdr** | **beta** | **p-value** | **fdr** | **beta** | **p-value** | **fdr** | **beta** | **p-value** | **fdr** | **beta** | **p-value** | **fdr** |
| **CryoAmount (millons per** | -0.010 | 0.423 | 0.634 | **-0.045** | **<0.001** | ***0.001*** | 0.005 | 0.637 | 0.845 | **0.056** | **<0.001** | ***######*** | **0.037** | **<0.001** | ***0.000*** | -0.003 | 0.825 | 0.983 | 0.001 | 0.913 | 0.998 | -0.011 | 0.386 | 0.634 | 0.026 | 0.070 | ***0.099*** | -0.013 | 0.200 | 0.451 |
| **Transfection efficiency** | 0.010 | 0.218 | 0.388 | 0.006 | 0.401 | 0.528 | -0.008 | 0.337 | 0.454 | **0.023** | *0.009* | ***0.040*** | -0.011 | 0.077 | 0.161 | -0.003 | 0.698 | 0.782 | **0.014** | *0.049* | 0.161 | 0.002 | 0.860 | 0.854 | **0.047** | **<0.001** | ***1.4E-05*** | 0.011 | 0.066 | 0.161 |
| **Cell viability** | -0.002 | 0.630 | 0.733 | 0.002 | 0.664 | 0.733 | 0.007 | 0.103 | 0.192 | -0.007 | 0.140 | 0.224 | 0.000 | 0.930 | 0.927 | **0.012** | *0.005* | ***0.023*** | **-0.011** | *0.007* | ***0.025*** | **-0.010** | *0.041* | 0.105 | -0.007 | 0.162 | 0.226 | **0.013** | **<0.001** | ***3.9E-04*** |

Three linear mixed-effect (LME) models were fit to explore relationships with measured DNA repair capacity: crude model; multivaraite model on individual level demographics; multivariate model for mixed technical-biological factors

Note: beta values presented here correspond to effect size of each variable by 1 unit increase.

Both the p values (p-value) from the original LME models and the multiple-testing corrected p values (fdr) are presented.

BMI: body mass index; CryoAmount: cell amount available for cryopreservation after isolation; fdr: false discovery rate.

**Supplementary Table 4.** Across-individual 10-pathway distances

| ID | 1 | 2 | 5 | 6 | 9 | 11 | 12 | 18 | 19 | 23 |
|---|---|---|---|---|---|---|---|---|---|---|
| 1 | 0.00 | 1.55 | 1.54 | 1.60 | 2.16 | 1.19 | 2.02 | 1.56 | 1.34 | 1.30 |
| 2 | 1.55 | 0.00 | 1.10 | 1.24 | 2.64 | 1.22 | 2.64 | 1.13 | 2.28 | 2.17 |
| 5 | 1.54 | 1.10 | 0.00 | 1.73 | 2.38 | 1.40 | 2.14 | 1.00 | 1.76 | 1.69 |
| 6 | 1.60 | 1.24 | 1.73 | 0.00 | 2.22 | 1.37 | 2.88 | 1.44 | 2.20 | 2.23 |
| 9 | 2.16 | 2.64 | 2.38 | 2.22 | 0.00 | 2.28 | 2.37 | 1.96 | 1.89 | 2.38 |
| 11 | 1.19 | 1.22 | 1.40 | 1.37 | 2.28 | 0.00 | 2.69 | 1.26 | 1.82 | 1.59 |
| 12 | 2.02 | 2.64 | 2.14 | 2.88 | 2.37 | 2.69 | 0.00 | 2.14 | 1.56 | 2.29 |
| 18 | 1.56 | 1.13 | 1.00 | 1.44 | 1.96 | 1.26 | 2.14 | 0.00 | 1.71 | 1.89 |
| 19 | 1.34 | 2.28 | 1.76 | 2.20 | 1.89 | 1.82 | 1.56 | 1.71 | 0.00 | 1.69 |
| 23 | 1.30 | 2.17 | 1.69 | 2.23 | 2.38 | 1.59 | 2.29 | 1.89 | 1.69 | 0.00 |

Note: across-individual distance in batch corrected data are calculated here (after taking mean of within individual measurements); average is 1.84.

**Supplementary Table 5.** Improved model fitting accuracy in repeated measures (CV%)

| ID | NLS pre | NLS post | Brms pre | Brms post |
|---|---|---|---|---|
| 1 | 31.2 | 12.0 | 20.0 | 4.7 |
| 2 | 82.7 | 41.5 | 38.4 | 34.4 |
| 5 | 59.8 | 27.1 | 44.7 | 18.4 |
| 6 | 28.1 | 41.0 | 29.5 | 40.0 |
| 9 | 37.4 | 21.6 | 27.7 | 19.6 |
| 11 | 26.8 | 20.9 | 22.4 | 19.0 |
| 12 | 40.0 | 20.8 | 34.5 | 19.5 |
| 18 | 50.5 | 37.7 | 39.8 | 27.6 |
| 19 | 74.4 | 31.8 | 37.2 | 22.0 |
| 23 | 64.3 | 31.5 | 37.6 | 25.7 |
| **average** | **49.5** | **28.6** | **33.2** | **23.1** |

NLS: non-linear square regression.

**Supplementary Table 6.** Individual differences in Comet kinetics

| Repair time | Intra-individual CV% | Inter-individual CV% |
|---|---:|---:|
| UT | 5.1 | 6.0 |
| 0min | 3.1 | 5.5 |
| 15min | 7.1 | 7.9 |
| 30min | 9.6 | 10.6 |
| 60min | 9.9 | 10.6 |
| 120min | 10.9 | 10.7 |
| t1/2 | 23.1 | 24.6 |
| t1/2 fast | 33.0 | 44.0 |
| t1/2 slow | 33.8 | 46.0 |

CV: coefficient of variation; UT: background damage level; t1/2: half life estimates of repair kinetics

**Supplementary Table 7**. Association between transfection efficiency (TE) and MGMT repair capacity.

| **Condition** | **beta** | **p-value** |
|---|---|---|
| Main analysis | 0.36 | 1.3E-06 |
| All samples, imputed with sample mean (285 obs) | 0.42 | 9.4E-06 |
| Complete samples (233 obs in 56 participants) | 0.59 | 5.7E-08 |
| Complete individuals (153 obs in 35 participants) | 0.45 | 2.0E-03 |

Note: Linear mixed-effects models for sensistivity analyses were adjusted for age, sex, BMI, smoking, race, cryoamount, cell viability, consistent with main analysis, with the only differences in batch correction method (main analysis used ComBat and sensitivity analysis used direct adjustment).
Beta values represent the change in MGMT repair capacity associated with an interquartile range (IQR) increase in TE, calculated by multiplying the model coefficient by the IQR of TE.

**Supplementary Table 8.** Interindividual variations in DNA repair capacity from previous literature

| Source Publication (DOI) | Repair | Sample Type | Sample Size | Mean | Standard Deviation | Fold Change | CV in Publication | CV in our assay |
|---|---|---|---|---|---|---|---|---|
| 10.1093/carcin/bgu214 | AAG | cell-free extracts | 97 | 161.00 | 34.00 | 3.3 | 21.12 | 15.6 |
| 10.1371/journal.pgen.1003413 | AAG | cell-free extracts | 80 | 0.46 | 0.24 | >10 | 52.03 | 15.6 |
| 10.1016/j.mrfmmm.2012.01.001 | AAG | cell-free extracts | 88 | 3.47 | 1.35 | 6.2 | 38.90 | 15.6 |
| 10.1016/s0921-8777(01)00096-9 | OGG1 | cell-free extracts | 34 | 33.05 | 5.88 | 2 | 17.79 | 9.5 |
| 10.1016/j.dnarep.2006.08.003 | OGG1 | cell-free extracts | 120 | 7.20 | 1.03 | 2.8 | 14.31 | 9.5 |
| 10.1158/1940-6207.CAPR-13-0318 | APE1 | PBMCs | 99 | 896.00 | 238.60 | 4.9 | 26.63 | 11.7 |
| 10.1093/carcin/4.12.1565 | UNG | liver | 2 | 2610.00 | 840.00 | 3.1 | 32.18 | 21.8 |
| | UNG | stomach | 2 | 1445.00 | 670.00 | 3.2 | 46.37 | 21.8 |
| | UNG | small intestine | 2 | 1640.00 | 1010.00 | 65 | 61.59 | 21.8 |
| | UNG | colon | 2 | 3170.00 | 1960.00 | 5.5 | 61.83 | 21.8 |
| | MGMT | liver | 2 | 1.07 | 0.62 | 7.8 | 57.94 | 79.8 |
| | MGMT | stomach | 2 | 0.20 | 0.04 | 1.7 | 20.00 | 79.8 |
| | MGMT | small intestine | 2 | 0.21 | 0.22 | 42 | 104.76 | 79.8 |
| | MGMT | colon | 2 | 0.14 | 0.08 | 9.6 | 57.14 | 79.8 |
| 10.1007/s002040100226 | MGMT | PBMCs | 40 | 105.00 | 56.00 | 7.6 | 53.33 | 79.8 |
| 10.18632/aging.100810 | APE1 | PBMCs | 18 | 50.93 | 29.81 | na | 58.53 | 11.7 |
| 10.1093/carcin/bgn140 | NHEJ | cell-free extracts | 93 | 29.60 | 10.70 | na | 36.15 | 8.8 |
| 10.1158/0008-5472.CAN-12-1915 | NER | PBMCs | 860 | 8.88 | 2.84 | 9.5 | 32.02 | 21.5 |
| | NER | PBMCs | 695 | 8.91 | 2.49 | 5.5 | 27.95 | 21.5 |

Note: full citations are provided in manuscript main text.

**Supplementary Table 9**. Plasmids for FM-HCR

| Cocktail | AmCyan | tagBFP | EGFP | mOrange | mPlum | Empty Vector |
|---|---|---|---|---|---|---|
| UD#1 | - | WT<br>100ng | WT<br>100ng | WT<br>100ng | WT<br>250ng | 8500ng |
| UD#2 | WT<br>2000ng | ScaI<br>100ng | pCNX-NNX/D3<br>500/5000ng | WT<br>100ng | C207G<br>500ng | 400ng |
| D#1 | - | **U:G** (A191G-U)<br>300ng | **Hx:T** (C289T-Hx)<br>100ng | **8oxoG:C** (A215C-8oxoG)<br>300ng | WT<br>250ng | 7650ng |
| D#2 | - | WT<br>100ng | **LP-BER** (617-THF)<br>100ng | **MMR** (G299C-G)<br>100ng | **A:8oxoG** (T202WT-8oxoG(+))<br>500ng | 7800ng |
| D#3 | WT<br>2000ng | **NHEJ** (ScaI linearized)<br>300ng | **HR** (D5G StuI linearized / D3)<br>500/5000ng | **NER** (UV 800 J/m2)<br>300ng | **MGMT** (T202C-C207G O6MeG)<br>500ng | - |

Note: plasmid reporters has been described in details in previous work cited in maintext (PNAS 2017, doi: 10.1073/pnas.1712032114). UD: undamaged; D: damaged.

### Supplementary Discussion

#### Technical factors influencing FM-HCR assay reproducibility in primary human PBMCs

Through a comprehensive study design with replicates and repeated measures spanning multiple batches, we explored key technical and biological factors influencing DRC measurements. Our findings identified batch effects as the primary source of variability in replicate measurements from the same individual and blood draw. This impact was observed across nearly all pathways, except direct reversal of O6MeG MGMT. Additionally, cryopreservation time was linked to variations in DRC but was ultimately explained by its joint effect with batch, as neither factor remained significantly associated with DRC after batch-effect correction. These results underscore the critical need to handle biological samples as consistently as possible and to account for batch effects when designing molecular epidemiological studies, for example by maximizing the number of samples included in each batch and minimizing inter-batch time gaps.

Beyond batch effects, we observed that the number of cells available at the time of cell preservation (CryoAmount**)** varied with individual demographics, while transfection efficiency and cell viability were influenced by both batch effects and demographic variables. These findings align with prior studies optimizing PBMC processing, which also highlighted donor variability as a key driver of technical variation [2,3]. Importantly, these factors affected DRC measurements even after batch-effect correction, adjustment for individual demographics, and multiple-testing correction, suggesting a combination of technical variability and inherent biological influences. Given the many links between DNA repair, immune function, and inflammation, it may be expected that differences in DRC are associated with the abundance, composition, and viability of blood cells [4-6] [7] [8].

High transfection efficiency could in principle affect DRC by leading to saturation or activation of the DNA damage machinery, or by increasing the representation of subpopulations of cells with inherently different repair capacity. Stimulated PBMCs exhibited high MGMT activity, with near-zero reporter expression on average (**Fig. 1**). In these cells, higher transfection efficiency was positively associated with increased MGMT repair capacity (est. = 0.36), a finding consistent across multiple sensitivity analyses (**Supplementary Table 7**). However, among PBMC samples showing maximal repair capacity (zero reporter expression), transfection efficiency tended to be lower, obscuring the trend and suggesting a misleading positive association between higher transfection efficiency and higher reporter expression (**Supplementary Fig. 6b**). The subtler increase in THF (LP-BER, est. = 0.17) repair with transfection efficiency was not investigated further, but could indicate that some damaged plasmids can trigger DNA damage response in PBMCs.

Although dead cells are excluded from our analysis [9], the percentage of viable cells may reflect aspects of sample handling and storage that could affect the phenotype of surviving cells. Interestingly, cell viability showed strong associations with NHEJ, HR, MMR, and NER, independent of batch noises and individual-level covariates

(**Fig. 4**). A common feature of the assays for measuring these DNA repair pathways is that the functional readout depends on multiple DNA repair proteins. For example, over two dozen proteins including the ~500 kDa THIIH complex are involved in the NER pathway and at least 10 lead to NER deficiency when mutated in humans. Thus, if any of these proteins is unstable under cryopreservation conditions and unable to fully recover its expression during the 72 hour incubation, repair efficiency could be impacted. By contrast, the readout of the reporter assays for BER and direct reversal of alkylation damage by MGMT are dominated by the activity of a single protein that is typically smaller than 50 kDa, and repair capacity as measured by these assays was not associated with cell viability.

PBMC viability can be affected by cell density during cryopreservation [10], thus the cells recovered from each blood draw were distributed equally into four vials regardless of the number of cells to minimize such impacts. Nonetheless, CryoAmount, the initial number of cells isolated from blood, varied considerably between individuals, indicating it is also likely a biological variable reflective of the number of circulating lymphocytes and potentially the immune function. CryoAmount was positively associated with repair of A:8oxoG and LP-BER, which may reflect a need for efficient BER during hematopoiesis and for stem cell resilience to DNA damage [11,12]. By contrast, an inverse relationship was found between CryoAmount and Hx:T, reminiscent of previous studies reporting that the AAG DNA glycosylase activity can be toxic in some contexts [13-16] or associated with increased lung cancer risk [17]. Future studies including more sophisticated immunophenotyping are needed to determine whether these observed associations reflect previously reported links between BER efficiency and immune cell populations [18].
